# The association between body mass index and metabolite response to a liquid mixed meal challenge

**DOI:** 10.1101/2023.08.21.23294369

**Authors:** David A. Hughes, Ruifang Li-Gao, Caroline J. Bull, Renée de Mutsert, Frits R. Rosendaal, Dennis O. Mook-Kanamori, Ko Willems van Dijk, Nicholas J. Timpson

## Abstract

**Background:** Metabolite abundance is a dynamic trait that is not only variable in a fasting state, but also varies in response to environmental stimuli, such as food consumption. Postprandial abundance and response to a meal are emergent traits in studies of disease and which themselves may be subject to specific risk factors. We investigated body mass index (BMI) as a recognized risk factor for numerous health outcomes that may influence metabolite response to feeding. Here we use the Netherlands Epidemiology of Obesity (NEO) study to examine associations between BMI and metabolite response to a liquid meal and extend this by using Mendelian randomization (MR) to estimate potential causal effects.

**Methods and findings:** The NEO study conducted a liquid meal challenge and collected metabolite profiles using the Nightingale metabolomics platform in 5744 study participants. Observational and one-sample MR analysis were conducted to estimate the effect of BMI on metabolites and ratios of metabolites (n = 229) in the fasting, postprandial and response (or change in abundance) states. After an appropriate multiple testing correction, we observed 473 associations with BMI (175 fasting, 188 postprandial, 110 response) in observational analyses. In MR analyses, we observed 20 metabolite traits (5 fasting, 12 postprandial, 3 response) to be associated with BMI. In both the fasting and postprandial state, this included citrate and the ratios of linoleic acid, omega-6 fatty acid and polyunsaturated fatty acids to total fatty acids. In addition, the glucogenic amino acid alanine was inversely associated with BMI in the response state, suggesting that as alanine increased in postprandial abundance, that increase was attenuated with increasing BMI.

**Conclusions:** Overall, MR estimates were strongly correlated with observational effect estimates suggesting that the broad associations seen between BMI and metabolite variation in fasting, postprandial and response states have a causal underpinning. Specific effects in previously unassessed postprandial and response states were detected and these may likely mark novel life course risk exposures driven by regular nutrition.

## Introduction

The excess accumulation of body fat and obesity are an established global health burden, the prevalence of which is increasing. The incidence and cost in life and dollars was estimated to be 1.9 billion cases in 2016, 4 million attributed deaths in 2015 and two trillion US dollars in 2014, respectively (1–3). Body mass index (BMI), or the ratio of one’s weight and height squared (kg/m^2^), is a common metric for measuring excess weight or fat and BMI thresholds of greater than or equal to a value of 25 and 30 are used to designate the conditions of overweight and obesity, respectively (4,5). While the use of BMI as an obesity metric – one that varies with age, sex, and ethnicity/genetic ancestry - may underestimate the true global burden of excess body fat, it has been an important and easily derived anthropometric and diagnostic metric in the study of obesity (6–9). Increases in BMI have been associated with decreased life expectancy (10–13), some cancers (14–16) and with cardiometabolic diseases including cardiovascular disease, and type 2 diabetes (17).

Despite the scale of the BMI related health burden, there remains a need to elucidate how and by what intermediate physiological traits excess body fat increases disease risk. Possible intermediates are metabolites and lipoproteins, some of which – like low density lipoprotein (LDL) or its’ crucial protein apolipoprotein B - have been previously associated with both BMI and disease risk (18–26). In a recall-by-genotype framework, previous work supports a causal role of body mass index on metabolite variation (27,28). Other work demonstrated a broad observational and consistent causal association between BMI and metabolites or specifically lipoprotein subclasses, branch-chain amino acids, and inflammatory markers in a young adult cohort (18).

In 1950, John Moreton illustrated the presence of intra-individual variability in chylomicrons after a high fat food (cream) challenge (29). More recent work has illustrated additional metabolite variation postprandially that could prove informative in evaluating disease risk and informing precision medicine (30–35). Yet, Schutte et al, building on the work of others (18), has illustrated that observational effect estimates between BMI and metabolite abundances are strongly, but not uniformly, consistent between fasted and postprandial metabolite abundance measurements (36). This observation suggests that for some metabolites fasting versus postprandial status may not be critically important for meta-analyses that aim to estimate the association between BMI and metabolite abundance. It also suggests that for other metabolites there is novelty in postprandial measurements and their association with BMI.

In this context and with most individuals spending much of their waking hours in a non-fasted or postprandial state, it is important to understand both the variation in postprandial metabolite abundance and the association and influence BMI may have on dynamic ranges in metabolomic response. Similar to an oral glucose tolerance test (OGTT), whose postprandial abundance is informative about an individuals’ insulin sensitivity and diabetic state, postprandial metabolite abundances may prove informative in the study of disease traits (37). Consequently, a comprehensive assessment of postprandial variation and response is essential in the study of adiposity, metabolites, and disease.

Response, defined as the change in metabolite abundance between fasted and postprandial states, has received little to no attention in disease risk research. In contrast to fasting status, response can be assessed either through a basic measurement of postprandial metabolite profile after a standardized feed (assuming that baseline variability is essentially random in a large enough sample), or through the explicitly modelling of response accounting appropriately for pre-feed variation. Either way, there is evidence that this dynamic state is different and potentially informative versus classically measured fasting levels. For example, a recent genome-wide association analysis identified genetic variants uniquely associated with metabolite response traits that did not share associations with fasting or postprandial abundances (38). Moreover, estimates of genotype heritability for some response traits such as the (branched-chain) amino acids, glucose and extremely large VLDL are as large or larger than those observed for fasting and postprandial estimates. Taken together, results suggest that such state specific traits are likely to be qualitatively different and may prove informative in explaining the aetiology of disease.

Currently missing from the literature are studies explicitly assessing the potentially causal relationship between variation in the commonly recognized risk factor BMI and the dynamic response of the metabolome to feeding. This is potentially difficult to examine given the strongly confounded nature of BMI as a risk factor, however Mendelian randomization (MR) is a framework using instrumental variable (IV) analysis to deploy genetic variants as exposure proxies in efforts to obtain evidence of causal relationships (39,40). It has been increasingly possible to exploit an increasing collection of confirmed genetic associations with BMI for the purpose of estimating likely downstream consequences in variation (41–49) and aggregate genetic scores have become useful in their capacity to capture heritable variance in exposures of interest (like BMI) despite potential complexities (50). There is, therefore, an opportunity to apply this approach to the examination of potential relationships between BMI and metabolite profiles.

We aimed to bring together data measuring the metabolic response to a liquid mixed meal and measures of the exposure, BMI, in observational and MR frameworks. These analyses, undertaken in the Netherlands Epidemiology of Obesity (NEO) study, sought to estimate relationships between variation in BMI and metabolite abundance in both a fasted and postprandial states, and to assess metabolite response – or change in abundance between a fasted and postprandial states.

## Materials and Methods

### Study Sample

The sample population was 6,671 participants of The Netherlands Epidemiology of Obesity (NEO) study, a prospective cohort study (51). The NEO study was conducted at the Leiden University Medical Center (LUMC) and approved by the LUMC Medical Ethical Committee. All participants gave written informed consent.

The NEO sample population is derived from two populations. First, all individuals between the ages of 45 and 65 years, living in the greater Leiden area, with a self-reported BMI of 27 kg/m^2^ or higher. Second, all individuals in the municipality of Leiderdorp of the same age range, regardless of BMI (**S1 Fig.**). We will refer to these two sample populations as Leiden and Leiderdorp respectively, and the entire study sample as NEO. Here our study sample population has a size of 5,744 defined by those participants who have genotype data, metabolite data, and other phenotype or covariable data available. Further details of the NEO study and study design are previously published (52).

### Meal challenge

An overnight fast was requested of all participants who subsequently visited the LUMC NEO study center and given a food challenge (38). During the visit an initial baseline, fasted blood sample was drawn. Second, within 5 minutes of the initial blood draw participants were given a liquid mixed meal. The meal was 400mL in volume with 600 kcal of energy. Sixteen percent of that energy (En%) was derived from protein, 50 En% from carbohydrates, and 34 En% was from fat. Third, following completion of the food challenge a blood sample was drawn at 30 minutes and again at 150 minutes. The NEO meal challenge occurred between the months of September 2008 and September 2012, with participants from Leiden sampled throughout this period of time. Those participants from Leiderdorp were sampled, non-exclusively, during the months of June 2011 and September 2012.

The meal challenge sampling of Leiderdorp and Leiden population participants was structured by sampling date and as a product BMI was structured by sampling date. Specifically, BMI was associated with the date of measurement (univariable linear model F-test p-value = 4.7×10^-229^), explaining 20.3% of the total variation in BMI (**S2 Fig.**). Despite this, the BMI-PGS had a smaller proportion (2.0%) of its variation explained by sampling date (univariate linear model F-test p-value = 3.9×10^-7^). As a product of this structure, sampling date is included as a covariate in all observational and MR analyses.

### Metabolite data

NEO participants had plasma derived metabolite profiles measured using the Nightingale Health (Helsinki, Finland) ^1^H nuclear magnetic resonance (NMR) platform (53). For each participant both their fasting and 150 minutes postprandial samples were assayed. We note here that metabolites are commonly defined as biological molecules less than 1.5 kilodaltons in size and that many of the molecules (lipids) assayed here are larger than this threshold. For simplicity we will refer to them all here as metabolites. At the time of sampling this metabolomics platform provided 229 measurements for 149 metabolites including 80 derived ratio measurements (**Table S1 in S1 File**) from 14 substance classes: amino acids (n=8), apolipoproteins (n=3), cholesterol (n=9), fatty acids (n=11), fatty acids ratios (n=8), fluid balance (n=2), glycerides and phospholipids (n=9), glycerides and phospholipid ratios (n=2), glycolysis related metabolites (n=3), inflammation (n=1), ketone bodies (n=2), lipoprotein particle size (n=3), lipoprotein subclasses (n=98), lipoprotein subclass ratios (n=70).

### Data quality control

Prior to data analysis, metabolites and covariable quality control and data filtering steps were implemented. First, a single individual who self-described themselves as “other” rather than “white” was excluded from the analysis. Second, any non-metabolite covariable with less than 1000 observations (n=1), with no variation (n=1), or binary covariables with fewer than 10 observations in either of the two binary classes (n=1) were removed. A complete list of evaluated non-metabolite covariables is available in **Table S2 in S1 File**.

The initial metabolite data set consisted of 5,744 individuals, 229 fasting metabolites, 229 postprandial metabolites, and 148 previously derived (38) orthogonal nonlinear least squares (ornls) response metabolite traits. Data quality control of this data set was performed with the R package *metaboprep* (54). A complete description of the procedure and parameters used with *metaboprep* can be found in **S1 Text.** After data QC 226 samples and 3 metabolites (all ornls response traits) were filtered from the data set (**S1 Log**). The log file (**S1 Log**) and report (**S1 Report**) generated by *metaboprep* are provided. After running *metaboprep* we also performed the following steps on the fasted and postprandial data. First, all zero values were converted into NAs. Second, for each metabolite (in the fasting and postprandial state, individually) all values 10 interquartile distances from the median were also turned into NAs. Third, we used the expected correlated nature of the fasting and postprandial data to identify outliers of that relationship, and turned them into NAs, in both dietary states. Further details on this procedure can be found in **S1 Text** and pre- and post-QC scatter plots for each metabolite can be found in **S2 File**.

### Response trait

A response trait, or a measure of change between the postprandial and fasting dietary state, was derived for each of the 229 metabolites traits by univariable Deming regression. A Deming regression was used here as it allows for error in both independent (fasting) and dependent (postprandial) variables of the model and as such is a type of total least squares regression. Following the QC steps described above we fit a Deming regression of postprandial on fasting data, for each metabolite. After model fitting the residuals were extracted and used as the response trait. The function deming() from the deming (v1.4) R package was used in the analysis. The intercept and slope of Deming regression were recorded and are provided in **Table S1** in **S1 File**, along with trait annotations and population summary statistics for all metabolite traits.

The residuals of the Deming regressions derived here were used as metabolite traits of response. These response traits effectively mirror those of a simple delta, in that positive values indicate an increase in metabolite abundance in the postprandial state and negative values would indicate a decrease. Effect estimates from an association analysis with BMI, described below, therefore suggests whether response is associated with BMI. Positive effect estimates suggest that as BMI increases, so too does the change in metabolite abundance.

Negative effect estimates suggest that as BMI increases the change in metabolite abundance decreases. Deming regressions residuals were taken to be the most appropriate way to summarize response for second stage analyses - observational and MR. However, in addition to the Deming regression, we also used two alternative methods to derive response. The first was a simple delta estimate (postprandial minus fasting abundance) and the second was an orthogonal nonlinear least squares (ornls) regression. We note that simple delta values and Deming residuals correlate with a mean Pearson’s r of 0.94 (95% CI 0.65-0.99). The ornls response traits were derived and used previously in a genome wide association study of metabolite response and was used in our metabolite quality control steps described above (38). Our result and discussion will focus exclusively on the Deming response traits, but all effect estimates for the delta and ornls response traits are provided in **S3 File**.

### Effective number of tested metabolites

Whilst there are 687 metabolite traits (fasting = 229, postprandial = 229, response = 229) these are not independent (**S3 Fig.**). We used the R package *iPVs* (https://github.com/hughesevoanth/iPVs) to estimate the effective number of metabolites in the data set (55–57). This allows us to ensure that we are not over correcting for the number tests we are performing in the study. We estimated 43 representative variables in the NEO metabolite data set. The data reduced, study-wide Bonferroni (BF) corrected p-value was set to 0.05/43 or 1.163×10^-3^. A full description of the iPVs procedure and parameters can be found in **S1 Text**.

### Metabolite data description

Two analyses were carried out to describe this Nightingale Health metabolomics data set. First, given the abundance of lipoproteins and their lipids in this data set we estimated the mean abundance of each lipid for each lipoprotein in both the fasting and postprandial dietary states. Second, a paired Student’s t-test was performed to determine if mean abundances differed between the fasting and postprandial states. Our threshold for declaring a change in mean abundance was 0.05/229, where 229 is the number of metabolites tested. We also estimated the change as a delta value between the postprandial and fasting states and then derived estimates for average change and the 95% confidence interval of the change distribution.

### Rank normal transformation of metabolite traits

Each metabolite trait distribution was tested for normality with a Shapiro-Wilk test. All W-statistics (untransformed distributions and model residuals) are reported in **S3 File**. In total 55.56% of all metabolite traits had a W-statistics less than 0.95, a threshold used here to define an inconsistency with normality. A total of 43.94% of all log-transformed metabolite traits had a W-statistics less than 0.95. Consequently, for the purpose of signal discovery and to allow for parametric analysis, each metabolite was rank normal transformed (tied values randomly ranked), prior to linear modelling. These steps resulted in the residuals from all observational and MR models to have a W-statistic greater than 0.9 and 0.99, respectively.

### Covariables

Variables from 14 categories were compiled and available to identify possible confounders by evaluating associations with BMI, BMI polygenic score (BMI-PGS), and metabolite traits (**Table S2** in **S1 File**). These 14 categories are: anthropomorphic (n = 8), education (n = 5), income (n = 2), smoking (n = 3), diet including alcohol intake in grams (n = 24), medication including glucose and lipid lowering (n = 10), systolic and diastolic blood pressure (n = 2), health including diabetes, glucose, hypertension and cancer status (n = 4), physical exercise (n = 18), basal metabolic rate (n = 1), indirect calorimetry (n = 5), genotype principal components (n = 4). In addition, seven metrics of biological sample quality (n = 7) and the sampling information (visit date and sub-population, n = 2) were included in the study to assess influence on metabolites. During quality control, described above, 3 variables were filtered. As performed with metabolites above we estimated the effect number of covariables in the data set. After running an iPVs analysis with a tree cut height of 0.5 we estimated that there are 54 representative or independent covariables. As a product any test performed across all covariables was corrected for multiple testing at a p-value of 0.05/54 or 9.29×10^-4^.

### Genotype data

The NEO participants were genotyped on the Illumina HumanCoreExome-24 BeadChip (Illumina Inc., San Diego, California, United States of America), at the Centre National de Génotypage (Evry Cedex, France). Following genotype quality control steps, as previously described (58), sample were imputed to the Haplotype Reference Consortium (HRC) release 1.1 (59), with further details in Li-Gao et al (38).

### Construction of the BMI polygenic score

A polygenic score for BMI (BMI-PGS) was derived for each individual using variants previously and independently associated with BMI (n=656) at a p-value less than 1×10^-8^ from the study by Yengo et al (60). To ensure the integrity of the PGS only those genetic variants, in the NEO genotype dosage data set, where the effect allele, the alternative allele, and the minor allele frequency (+/- 0.1) matched the data from Yengo et al were retained. If a match could not be made, then that SNP was dropped. All effect estimates were aligned to be positive, such that the effect allele was always that which increased an individual’s genotype predicted BMI. Finally, a weighted PGS was constructed by weighting the number of BMI increasing alleles, at each SNP (n=646, after data harmonization), by the effect estimate for that SNP and then summing across all values (60). The R script “01_generate_bmi_grs.R” for deriving the PGS can be found in the github repository https://github.com/hughesevoanth/NEO_BMI_Metabolite_MR.

### Observational and Mendelian randomization analysis

Two observational association analyses were undertaken in cross-section. The first was an association analysis between individual metabolite traits and BMI. The second uses a PGS as a proxy instrument for the exposure in a Mendelian randomization framework to provide an estimate of the causal effect of the exposure on outcome (here dietary state specific metabolite values). We will refer to the first as an observational analysis and the later as an MR analysis.

Observational analyses were performed by a multivariable generalized linear regression with sampling data, sub-population, age, and sex as additional covariables - taking the form of glm(metabolite ∼ visit date + sub-population + age + sex + BMI). For each generalized linear model we also perform a Breusch-Pagan test of homoskedasticity using the bptest() function from the lmtest R package (61). In addition, for each association analysis we performed a Type I analysis of variance or ANOVA producing a table of deviances and estimated an eta-squared statistic (17^2^), providing an estimate of the variance explained, for the model and for the primary exposure, BMI (**S3 File**).

One-sample Mendelian randomization analyses were performed using the ivreg R package (https://cran.r-project.org/web/packages/ivreg/), which implements a two-stage least squares instrumental variable analysis. In all instances and given the relative viability of BMI instruments versus reciprocal metabolite instrumentation, BMI was defined as our exposure, the PGS described above is our instrument for BMI and each metabolite trait was iteratively defined as the outcome. The same model and covariables as described in the association analysis paragraph above was used here. Along with MR effect estimates, we also report the Breusch-Pagan test of homoskedasticity, the F-statistic and F-test p-value testing for weak instruments, and summary statistics for the Durbin-Wu-Hausman endogeneity test (**S3 File**).

Unless stated otherwise - because our outcomes are rank normal transformed (zero centered with a standard deviation of one) - the effect estimates (beta) and standard errors (se) are reported as rank normalized standard deviation units change per unit increase in BMI in kilograms per meter-squared (kg/m^2^).

### Study structure

In all observational and MR analyses, four (sub-)population analyses were performed. The primary analysis, in all instances, was a weighted NEO (wNEO) analysis using data from all available samples. The wNEO analyses included weights for each sample that were previously derived to have the Leiden sub-population BMI distribution emulate that of the randomly sampled Leiderdorp sub-population (62). Sensitivity analyses include (1) a Leiderdorp sub-population analysis, (2) an un-weighted NEO analysis, and (3) a Leiden sub-population analysis. In addition, sex specific analyses were carried out using the primary weighted NEO framework and the Leiderdorp sub-population, as neither have a sampling bias in there BMI distributions.

### Code availability

All statistical analysis were conducted in the R language (v 4.0.2, Taking Off Again), and all bespoke functions and analytical code for the study can be found in the github repository https://github.com/hughesevoanth/NEO_BMI_Metabolite_MR.

## Results

### NEO cohort description

Following quality control and derivation of the response trait the data set consisted of 5517 individuals (51.6% female), 687 metabolite traits (229 fasting, 229 postprandial, 229 response) and 85 covariables. The average age of participants was 56 years, with an average BMI of 29.98 kg/m^2^ (**Table 1**).

**Table 1.** Population summary statistics. Population summary statistics for the (1) complete NEO cohort, for (2) men and (3) women of the NEO cohort and the two NEO sub-populations (4) Leiderdorp and (5) Leiden, respectively. For each variable: age, height, weight, waist circumference, hip circumference, waist-hip ratio, smoking in pack years, alcohol consumption in grams per day, the percent of the population with higher education, BMI, and BMI-PGS the mean (95% confidence intervals) is provided. In addition, the number of men and women in each sample (sub-)population is provided (sex). The last three rows of data provide summary statistics describing the relationship between the instrumental variable (BMI-PGS) on the exposure (BMI). Specifically, the variance explained (R^2^), the effect estimate (beta) and standard error (se), and the P-value (P) are provided. Estimates suffixed with ** are derived from linear models that include sample weights. The last two columns of data provide an estimate of the proportion of variation (in NEO) of the trait (in the row) explained by variation between the sub-population samples, as estimated by a linear regression, and the model P-value.

| Trait | NEO | Men | Women | Leiderdorp | Leiden | var exp<br>by sub-<br>pop | p-value |
| --- | --- | --- | --- | --- | --- | --- | --- |
| sex | M:2671<br>F:2846 | M:2671<br>F:0 | M:0<br>F:2846 | M:623<br>F:783 | M:2048<br>F:2063 | 0.17% | 3.65E-04 |
| age | 55.99<br>(45.00-65.00) | 56.13<br>(45.00-65.00) | 55.86<br>(45.00-65.00) | 56.13<br>(46.00-65.00) | 55.95<br>(45.00-65.00) | 0.02% | 3.26E-01 |
| height | 1.74<br>(1.57-1.92) | 1.81<br>(1.68-1.95) | 1.67<br>(1.55-1.79) | 1.73<br>(1.57-1.92) | 1.74<br>(1.57-1.92) | 0.03% | 2.03E-01 |
| weight | 90.58<br>(60.00-126.80) | 97.45<br>(72.95-131.60) | 84.14<br>(57.20-121.52) | 79.16<br>(54.00-116.15) | 94.49<br>(69.60-128.85) | 16.12% | 9.63E-213 |
| waist<br>circ. | 102.01<br>(74.00-129.00) | 106.32<br>(85.79-131.00) | 97.96<br>(71.00-126.00) | 91.22<br>(70.00-119.00) | 105.70<br>(86.00-131.00) | 22.87% | 2.85e-313 |
| hip<br>circ. | 110.23<br>(94.00-133.00) | 108.57<br>(96.00-125.00) | 111.79<br>(92.00-138.00) | 103.40<br>(90.00-124.17) | 112.57<br>(99.00-135.00) | 16.22% | 3.33E-214 |
| pack<br>years | 11.21<br>(0.00-52.38) | 13.71<br>(0.00-61.69) | 8.88<br>(0.00-43.83) | 8.69<br>(0.00-43.32) | 12.09<br>(0.00-55.20) | 0.83% | 1.01E-10 |
| alcohol | 15.52<br>(0.00-60.52) | 21.40<br>(0.00-73.24) | 10.00<br>(0.00-41.72) | 14.58<br>(0.00-54.06) | 15.84<br>(0.00-62.57) | 0.10% | 1.95E-02 |
| smokin<br>g<br>(N\|F\|C) | 34.11 \|<br>49.85 \| 16.03 | 30.32 \|<br>50.49 \| 19.19 | 37.67 \|<br>49.26 \| 13.07 | 39.29 \|<br>46.76 \| 13.95 | 32.34 \|<br>50.91 \| 16.74 | 0.38% | 4.11E-06 |
| higher<br>ed % | 38.58 | 42.27 | 35.12 | 50.5 | 34.48 | 1.52% | 6.24E-26 |
| BMI | 29.98<br>(21.40-41.35) | 29.74<br>(22.72-38.99) | 30.20 (20.64-42.90) | 26.24<br>(19.94-37.20) | 31.26<br>(26.02-42.19) | 20.70% | 3.66E-280 |
| BMI-PGS | 10.20<br>(9.64-10.78) | 10.20<br>(9.63-10.78) | 10.20<br>(9.65-10.77) | 10.14<br>(9.56-10.71) | 10.22<br>(9.67-10.80) | 1.63% | 1.53E-21 |
| R<sup>2</sup> | 0.0431,<br>0.0473** | 0.0439 | 0.0453 | 0.0427 | 0.0243 | - | - |
| beta<br>(se) | 3.38 (0.21),<br>3.26 (0.20)** | 2.77 (0.25) | 4.00 (0.34) | 3.14 (0.40) | 2.25 (0.22) | - | - |
| P | 8.10x10<sup>-55</sup><br>3.61x10<sup>-60**</sup> | 7.23x10<sup>-28</sup> | 1.57x10<sup>-30</sup> | 5.10x10<sup>-15</sup> | 8.22x10<sup>-24</sup> | - | - |

The NEO cohort is made up of two distinct sub-samples, (1: “Leiderdorp”) a randomly sampled population of 1,406 individuals from the municipality of Leiderdorp (mean BMI = 26.24 kg/m^2^, 95% quantile interval (QI)19.94-37.2), and (2: “Leiden”) a sample population of 4,111 participant from the city of Leiden who were oversampled for individuals with a BMI greater than 27 kg/m^2^ (mean BMI = 31.26 kg/m^2^, 95% QI 26.02-42.18). BMI distributions differed between the two sub-populations (generalized linear model (glm) Student’s t test p-value = 3.66×10^-280^, **S1 Fig.**). In addition, the two sub-populations also differ in sex ratio, alcohol intake (g/day), smoking (packyears), educational attainment and BMI polygenic score (BMI-PGS; generalized linear model Student’s t test p-value < 1.95×10^-2^), but not in age or height (**Table 1**). Whilst the variance explained by sub-populations is large for BMI (20.7%) and closely related traits (weight, hip and waist circumference), it is an order smaller for other variables like BMI-PGS, packyears, and educational attainment (variance explained by sub-population < 2%, **Table 1**).

### Description of metabolite data

Of the 229 assayed metabolites 98 are lipoproteins and their particle concentrations and an additional 70 are lipoprotein ratios. The expected lipid concentrations were observed across lipoprotein classes (**S4 Fig.**). Namely, triglycerides dominate VLDLs, cholesterol dominates IDL and LDLs, and phospholipids and cholesterol dominate HDLs, on average. This remains true in both the fasting and postprandial data. All bar 14 metabolites differed in mean concentration or ratio between the fasted and postprandial states (paired t-test, P < 0.05/229, **Table S3** in **S1 File**). These 14 include the concentration of particles: total lipids, total cholesterol, cholesterol esters, and free cholesterol in medium LDL. It also includes total cholesterol in LDL, large LDL, and small LDL, and cholesterol esters in small LDL. Overall, 126 metabolites increased in abundance, and 89 decreased postprandially (**Table S3** in **S1 File**). Those with the largest scaled and centred, mean increase were the amino acids tyrosine, leucine, valine, isoleucine, and phenylalanine (**S5 Fig.**). Those with the largest mean decrease were the ratio of saturated fatty acids to total fatty acids, the ratio of triglycerides to total lipids in very large VLDL, the ratio of phospholipids to total lipids in small VLDL and the ketone body 3-hydroxybutyrate **(Table S3** in **S1 File**). For all metabolites, 150-minute postprandial change was not in the same direction for all participants (**S5 Fig.**).

### The observational effect of BMI on metabolite trait variation

wNEO observational analyses suggested broad association between BMI and metabolite traits, with 473 metabolite traits (across fasting, postprandial, and response states) showing evidence of association with BMI (P < 1.163×10^-3^, **Fig. 2, S6 Fig, and S3 File**). Effect estimates correlated between sexes (Pearson’s r = 0.965), but more associations were observed in women (426 total: 162 fasting, 175 postprandial, 89 response) than men (369 total: 161 fasting, 168 postprandial, 40 response, **Fig. 3A**). A total of 333 associations were shared between men and women, with 93 specific to women, 36 specific to men, and 225 metabolite traits having no association with BMI in either sex (**S3 File**). Among the associated traits all but 10 are directionally consistent among men and women (**Fig. 3A**). Larger effect sizes were observed in men (mean absolute beta = 0.0356) than women (mean beta = 0.0291), across all traits on average (**Fig. 3A)**, but the variance explained by BMI, across all traits on average was similar among men and women (η^2^: men = 1.737%, women = 1.734%).

**Figure 1.**
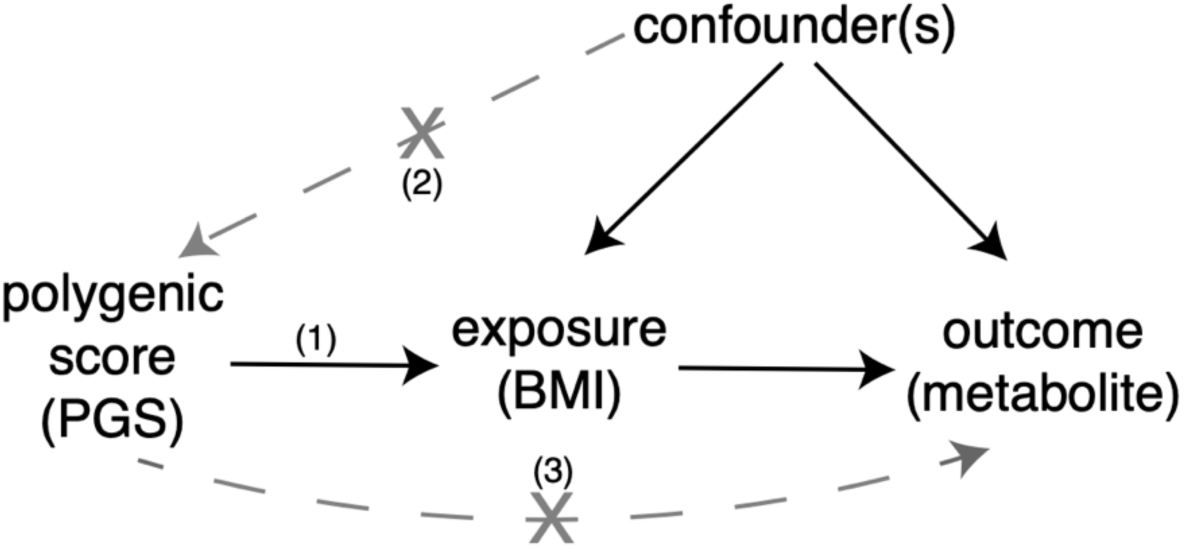
Mendelian Randomization schematic. A schematic or directed acyclic graph of the Mendelian randomization framework. The MR framework in this study assumes (1) that the instrument is robustly associated with the outcome, (2) that the instrument is not associated with variables that are associated to the outcome and (3) that the instrument acts on the outcome only through the exposure and not through a pleiotropic pathway.

**Figure 2:**
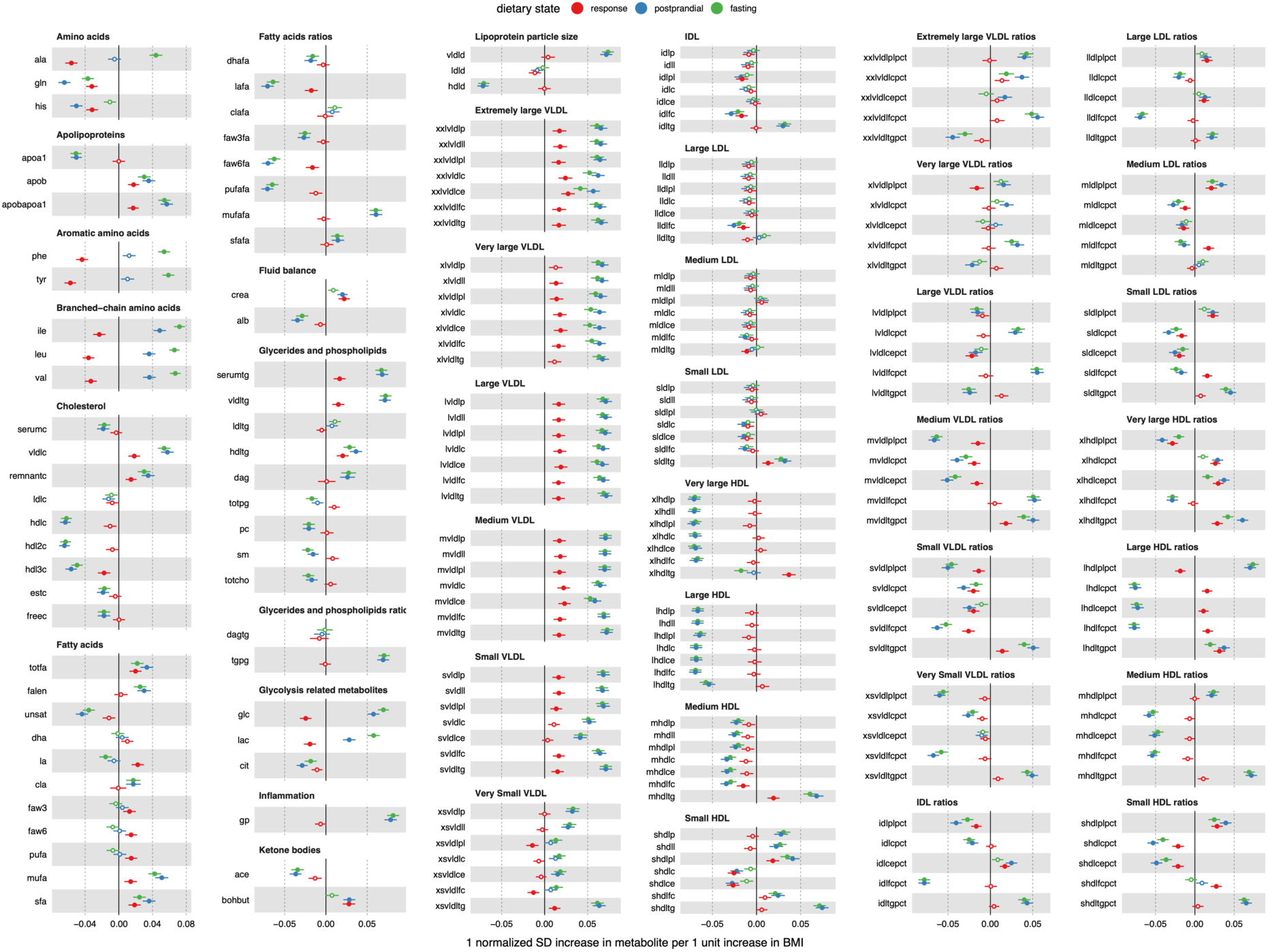
Observational BMI – metabolite abundance and response effect estimates. Point estimates and 95% confidence intervals for BMI on fasting metabolite abundance (green), postprandial metabolite abundance (blue), and metabolite response (red). Metabolites are classified and organized by Nightingale Health sub-class assignments. Metabolites that are associated with BMI, after multiple test correction (P<0.05/43), are indicated as solid color point estimates.

**Figure 3:**
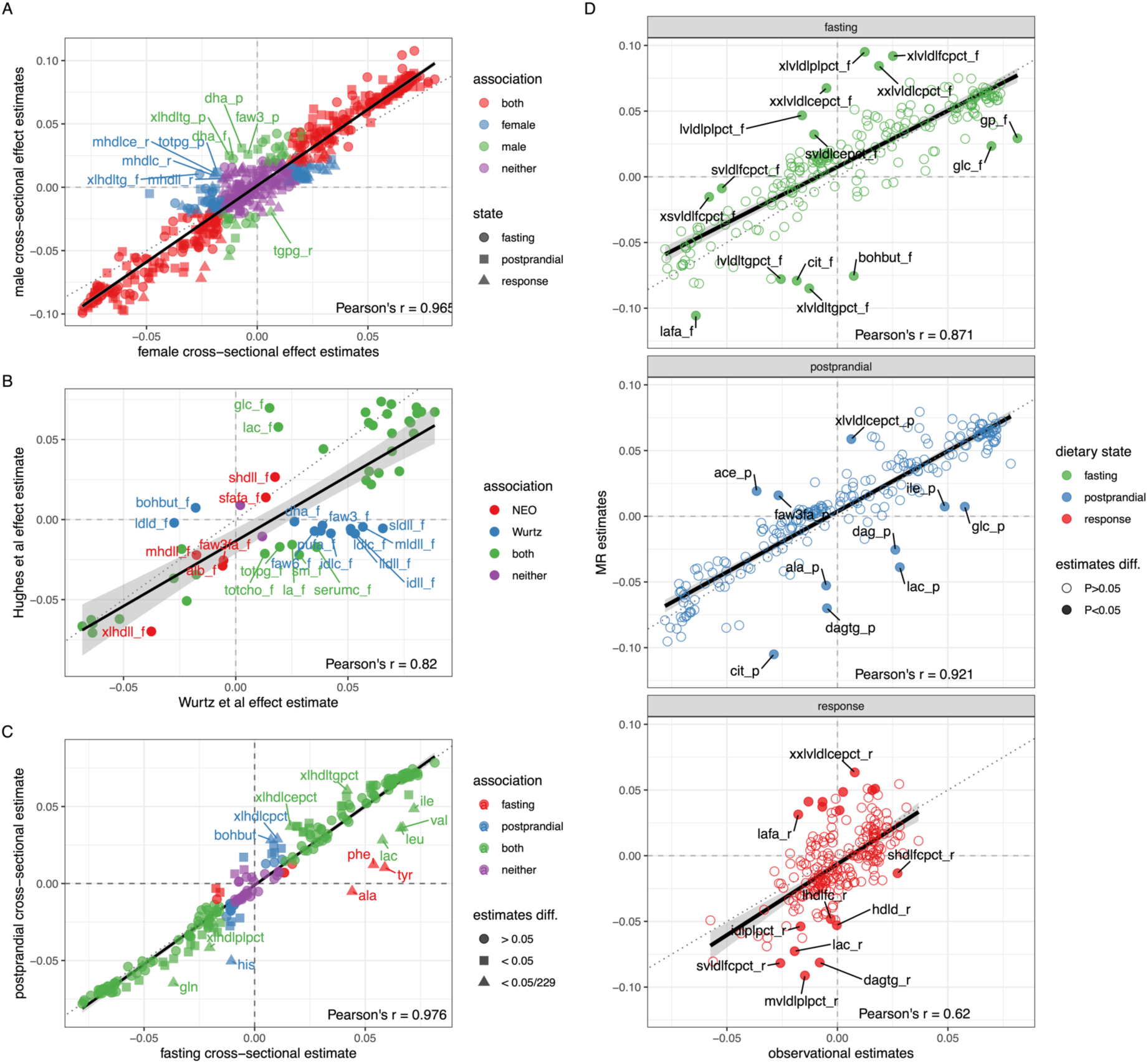
Scatter plots. In all plots the solid black line is a best fit regression line, the dotted black line is an equivalency line (intercept = 0, slope = 1), the dashed vertical and horizontal line indicate the zero values, and a Pearson’s correlation coefficient is in the bottom right corner**. (**A) Scatter plot of BMI-metabolite cross-sectional effect estimates for females (x-axis) and males (y-axis). Point estimates are colored red if there is an association between the metabolite and BMI in both sexes, blue if an association was only observed in females, green if an association was only observed in males, and purple if no association was observed in either sex. Point estimates are shaped as circles, squares, and triangles to represent the three dietary states fasting, postprandial, and response, respectively. (B) Scatter plot of BMI-metabolite cross-sectional effect estimates from Wurtz et al (x-axis) and NEO (this study, y-axis). Point estimates are colored red if there is an association between the metabolite and BMI in NEO, blue if an association was only observed in Wurtz et al, green if an association was observed in both studies, and purple if no association was observed in either study. (C) Scatter plot of BMI-metabolite cross-sectional effect estimates for fasting (x-axis) and postprandial (y-axis) dietary states. Point estimates are colored red if there is an association between the metabolite and BMI in the fasting state, blue if an association was only observed in the postprandial state, green an association was observed in both dietary states, and purple if no association was observed in either. Point estimates are shaped as circles if they are not different between the two dietary states – as determined by a z-test – shaped as squares if they are nominally different (P < 0.05) and triangles and labeled if they are different after correcting for multiple tests (P < 0.05/229). (D) Scatter plot of BMI-metabolite cross-sectional (x-axis) and MR (y-axis) effect in the fasting (top and green), postprandial (middle and blue), and response (bottom and red) dietary states. Metabolites with effect estimates that differ between the cross-sectional and MR analyses are solid circles and labelled with the metabolite name.

### Observational results - fasting

A total of 175 or 76% of fasting traits showed evidence of association with BMI. The strongest association observed was with the inflammation marker glycoprotein acetyls (beta = 0.081, se = 0.0037, P = 2.89×10^-101^). This was followed closely by the branched chain amino acids isoleucine (beta = 0.072, se = 0.0033, P = 4.18×10^-101^), leucine (beta = 0.066, se = 0.0031, P = 1.16×10^-94^) and valine (beta = 0.067, se = 0.0032, P = 8.70×10^-92^). The strongest inverse associations were observed for average diameter of HDL particles (beta = -0.071, se = 0.0035, P = 6.74×10^-95^), the ratio of free cholesterol to total lipids in IDL (beta = -0.078, se = 0.0036, P = 1.04×10^-100^) and large HDL (beta = -0.077, se = 0.0037, P = 8.09×10^-95^). If data is summarized by metabolite annotation class (**Table S1** in **S1 File**), we find that the classes with the largest average absolute BMI effect are inflammation (average absolute beta = 0.081), amino acids (0.051), lipoprotein particle size (0.049) and glycolysis related metabolites (0.049). Those with the smallest average absolute effect are ketone bodies (0.021), fluid balance (0.019) and fatty acids (0.018) (**S6 Fig.**).

### Observational results - postprandial

A total of 188 or 82% of postprandial traits showed evidence of association with BMI. Fasting and postprandial point estimates correlate strongly (Pearson’s r = 0.976) with each other, with no mean difference in their distributions (t-test P = 0.83) and 166 shared associations. However, 31 associations are unique to one of the two dietary states, with nine associations specific to fasting and 22 specific to postprandial data (**Fig. 3C**). We tested for a difference in effect estimates between the dietary states and observed 14 estimates to differ (z-test, P<0.05/229, **Fig. 3C**). Eight of those 14 are amino acids (tyrosine, alanine, phenylalanine, histidine, valine, leucine, glutamine, and isoleucine). The other six that differ between dietary states include four very large HDL ratios, the ketone beta-hydroxybutyrate (bohbut) and the glycolysis related metabolite lactate (lac).

### Observational results - response

A total of 70 response traits showed evidence of a positive association with BMI and 40 traits had an inverse association. Response and fasting (Pearson’s r = 0.34, P = 1.39×10^-7^), and response and postprandial (Pearson’s r = 0.514, P = 7.96×10^-17^) point estimates modestly correlate with each other, indicative of the relative independence of the response trait. The strongest associations were all amino acids, which (ordered by association P-value: tyrosine, alanine, phenylalanine, leucine valine, glutamine, histidine, and leucine) were all inversely associated with BMI (**S6 Fig.**). For example, while alanine levels (fasting mean = 0.359 mmol/L) were elevated after a liquid mixed meal (postprandial mean = 0.376 mmol/L, paired t-test P = 2.53×10^-181^) the effect attenuated as BMI increased (beta = -0.056, se = 0.004, P = 5.72×10^-51^, **Fig. 4a**) such that the effect of BMI on alanine response was negative (**Fig. 4b**). This relationship was observed for all amino acids in the data set (**Fig. 2**).

**Figure 4.**
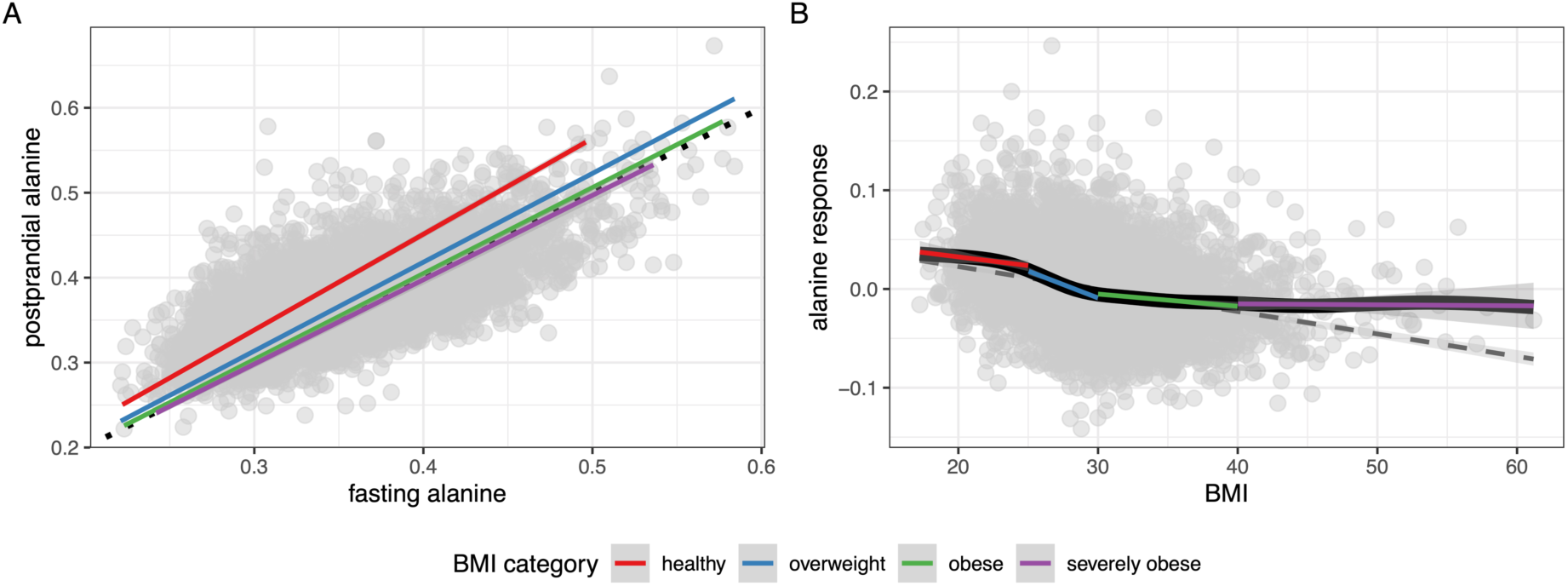
Association of BMI with alanine response. Plot on the left illustrates the correlation between fasting (x-axis) and postprandial (y-axis) tyrosine abundance, with a best fit line for 4 BMI classes of individuals: healthy (BMI < 25), overweight (25 <= BMI < 30), obese (30 <= BMI < 40), and severely obese (BMI > 40). All best fit lines were forced through an intercept of zero. Note that the slope of the best fit line is largest for individuals who have a healthy BMI and smallest for those who have severe obesity. The plot on the right illustrates the inverse relationship between BMI (x-axis) and tyrosine response (y-axis). The blue line is a best fit linear regression line, the black line is a best fit generalized additive model (GAM) smooth (non-linear regression), and the red, blue, green, and purple line intervals are best fit regression for each clinically defined portion of the BMI distribution, which mirror the non-linear regression.

The strongest positive response association was observed for triglycerides in very large HDL (xlhdltg: beta = 0.037, se = 0.004, P = 5.88×10^-21^). This was followed closely by the ratio of (1) cholesterol esters, (2) triglycerides, and (3) total cholesterol to total lipids in very large HDL. Conversely, the response trait ratio of phospholipids to total lipids in very large HDL decreased as BMI increased. This suggests that following a liquid mixed meal the increase of phospholipids relative to total lipids in very large HDL is attenuated by BMI and conversely, the ratio of cholesterol and triglycerides to total lipids in very large HDL increased as BMI increased (**Fig. 2**).

Outside of amino acids and lipoproteins, other metabolite response traits that had a positive association with BMI included 3-hydroxybutyrate (ketone body), creatinine (fluid balance), linoleic acid, and total fatty acids (fatty acids). Others with an inverse association with BMI included glucose and lactate (glycolysis), the ratio of linoleic acid to total fatty acids, and the ratio of omega-6 fatty acids to total fatty acids (fatty acid ratios; **S3 File**).

### Possible confounders

The instrumental variable used, BMI-PGS, explains 4.7% of the variation in BMI, and for each unit increase in BMI-PGS there is a 3.26 (se = 0.20) unit (kg/m^2^) increase in BMI (P = 3.61×10^-60^, wNEO analysis, **Table 1**). An assessment of the correlation between BMI and BMI-PGS with study covariables was performed to identify possible confounders, specifically those that would violate the second MR assumption (exchangeability). Of the 91 tested covariables, 60 were associated with BMI (wNEO, P < 9.26×10^-4^; **Table S4** in **S1 File** and **S7 Fig.**), and 17 were associated with BMI-PGS (**S8 Fig.**). Those 17 that associated with BMI-PGS included other adiposity traits (n = 5), sub-population, visit date, genetic principal component 3, (imputed) smoking packyears, on a weight loss diet, type of diet, on a diet last month, a food rule, mean CO_2_ production, and a peroxidation sample quality flag. The 17 covariables associated the BMI-PGS were tested for an association with each metabolite trait (the outcomes) in a univariable linear model. Results illustrated broad association across all traits and covariables, defining each covariable as a confounder in MR analysis leading to sensitivity analyses discussed below (**S9 Fig.**). A more comprehensive description of these analyses is in **S1 Text**.

### Estimating the causal effect of BMI on metabolite trait variation

MR analyses (wNEO) support broad associations between BMI and metabolites as suggested by the overall strong correlation between observational and MR estimates (Pearson’s r = 0.852, P = 1.02×10^-194^, intercept = 1.54×10^-03^, slope = 0.911). The correlation was strongest for postprandial traits (Pearson’s r = 0.921, P = 4.44×10^-95^, intercept = 3.59×10^-03^, slope = 0.922) and weaker for fasting (Pearson’s r = 0.871, P = 7.71×10^-72^, intercept = 7.62×10^-03^, slope = 0.855) and response traits (Pearson’s r = 0.62, P = 1.11×10^-25^, intercept = -6.36^-03^, slope = 1.08, **Figure 3D**). Forty-two observational and MR effect estimates showed evidence of being different from each other (z-test P< 0.05), yet none remain so after correcting for multiple tests (P < 0.05/687). Overall, these observations indicate that cross-sectional estimates have reasonable power at predicting MR estimates for these exposure-outcome analyses, consistent with previous work (18).

A total of 201 metabolite traits (across fasting, postprandial, and response states; **Figure 5**, **S3 File**) showed nominal evidence of association (P < 0.05) with BMI and 20 (**Table 2**, **Figure 6**) showed evidence of association with BMI (P < 1.16×10^-3^; **S10 Fig.**). MR effect estimates varied more between sexes (Pearson’s r = 0.368, p = 1.93×10^-23^) than observed in observational results, with 86 showing evidence of being different from each other (z-test P< 0.05), yet none remaining so after correcting for multiple tests (P < 0.05/687). Nine metabolite traits (7 fasting, 2 response) showed evidence of association with BMI in females, eight of which were unique to females (**Figure 6**). This included fasting small HDL triglycerides (shdltg_f) and fasting very small VLDL triglycerides (xsvldltg_f), both of which increased with increases in BMI. In contrast, five metabolites were associated with BMI in men, one of which was unique to men – postprandial glutamine, which had an inverse relationship with BMI (gln_p; **Figure 6**). The other four metabolites shared an association with the general population. They are, fasting and postprandial citrate (cit_f, cit_p), and the ratio of linoleic acid to total fatty acids in both the fasting and postprandial state (lafa_f, lafa_p). Each of which were also inversely associated with BMI. On average MR effect estimates were larger in females (absolute mean beta = 0.047) than males (absolute mean beta = 0.026, paired t-test P = 1.43×10^-45^).

**Figure 5:**
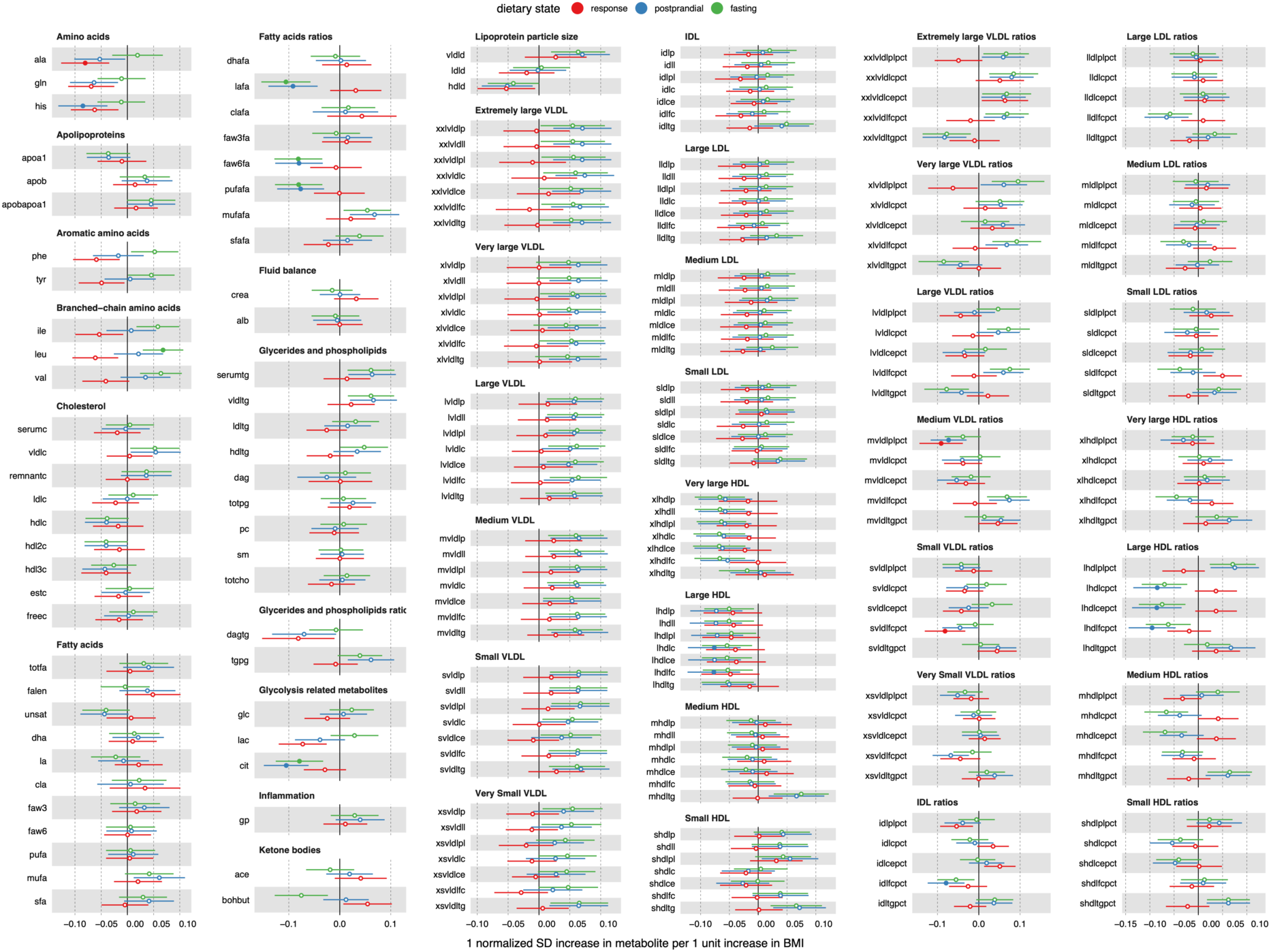
Forest Plot of MR effect estimates. Point estimates and 95% confidence intervals for BMI on fasting metabolite abundance (green), postprandial metabolite abundance (blue), and metabolite response (red). Metabolites are classified and organized by Nightingale Health sub-class assignments. Metabolites that are associated with BMI, after multiple test correction (P<0.05/43), are indicated as solid color point estimates.

**Figure 6:**
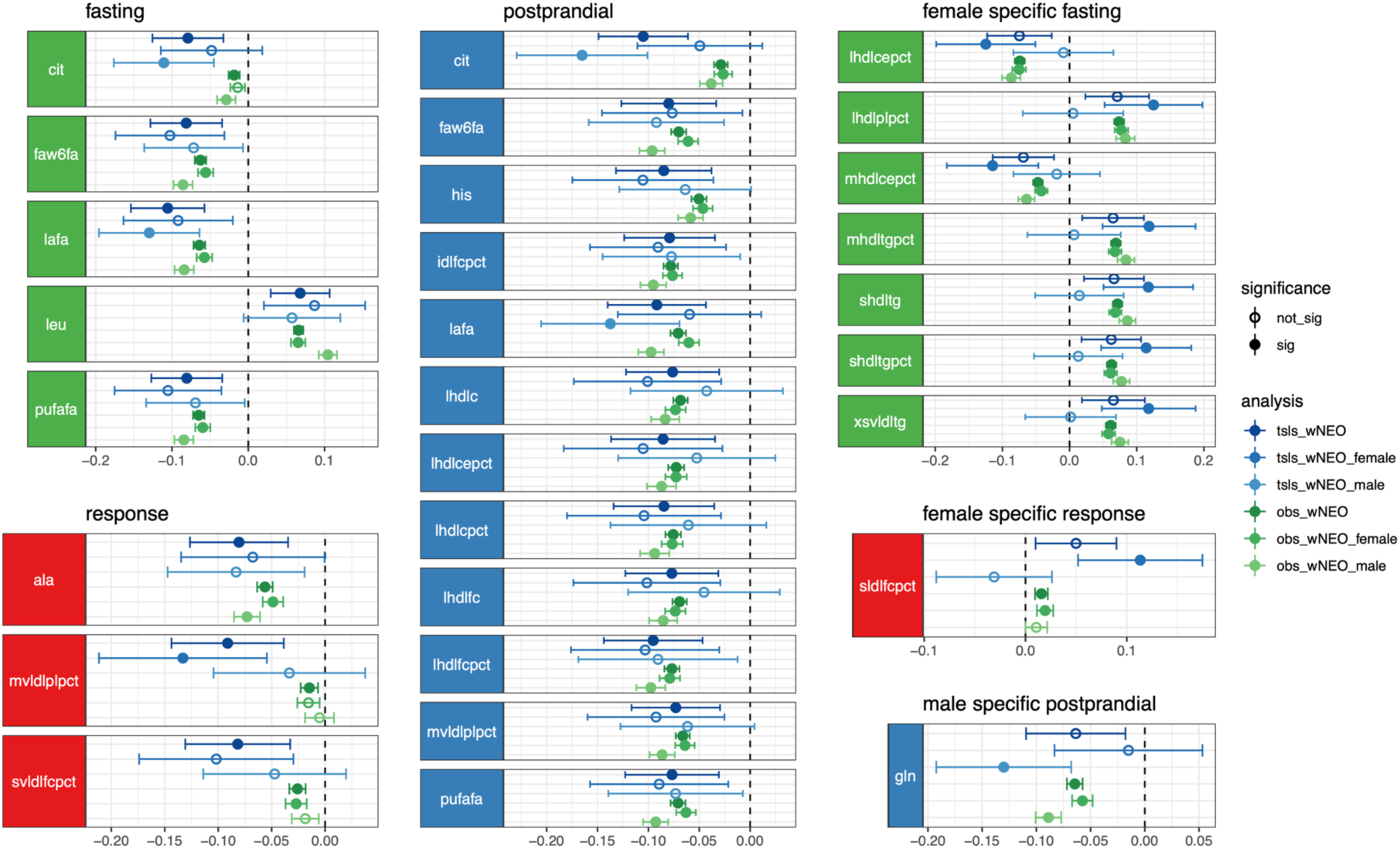
Metabolites associated with BMI in MR analyses. A forest plot of effect estimates (points) and 95% confidence intervals (whiskers) for MR (blue, “tsls” prefix) and observational (green, “obs” prefix) effect estimates for the weighted NEO (wNEO), weighted NEO female (wNEO_female), and weighted NEO male (wNEO_male) (sub-)populations. Fasting, postprandial, and response association observed in the general population as well as those association only seen in females and males.

**Table 2.**
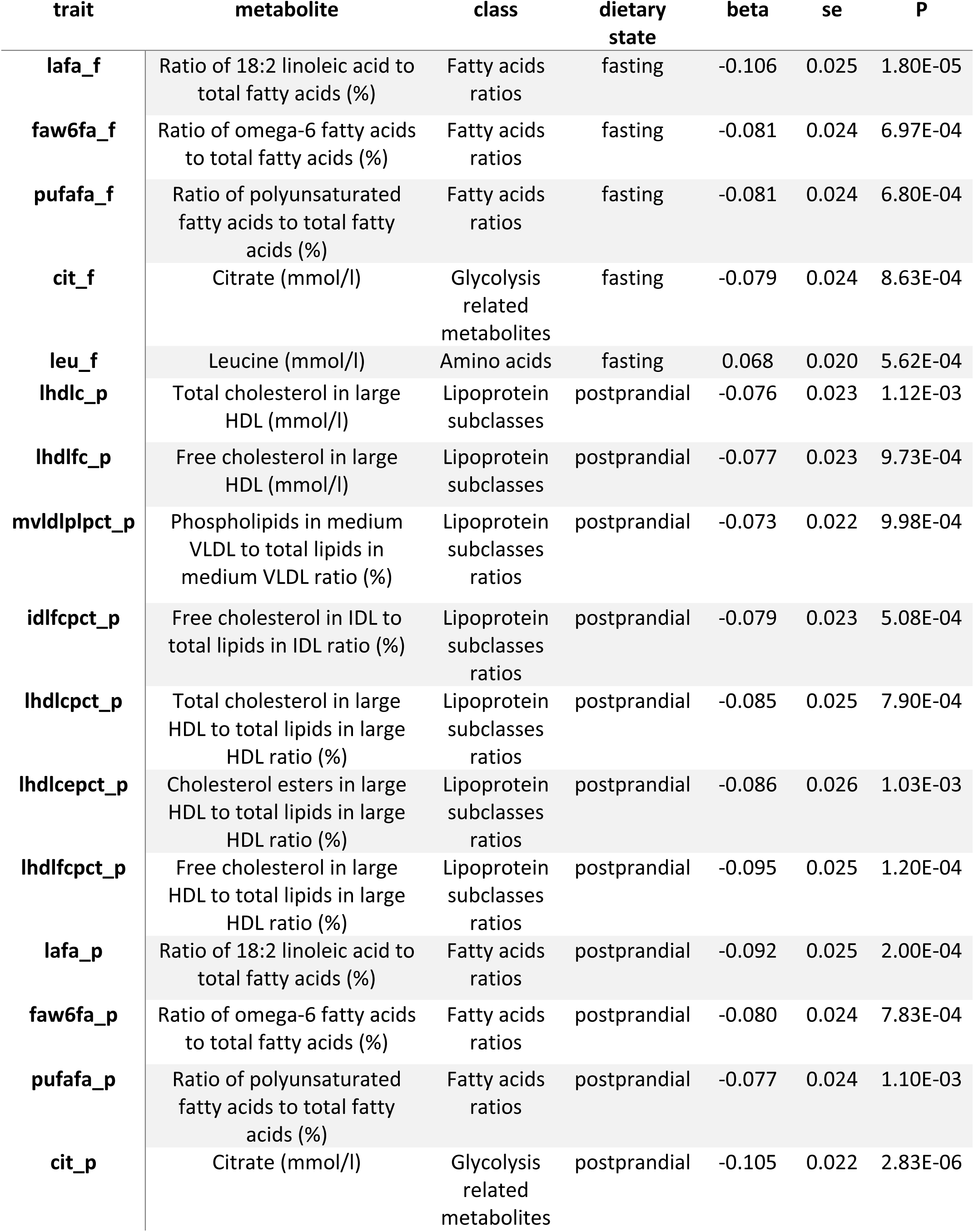
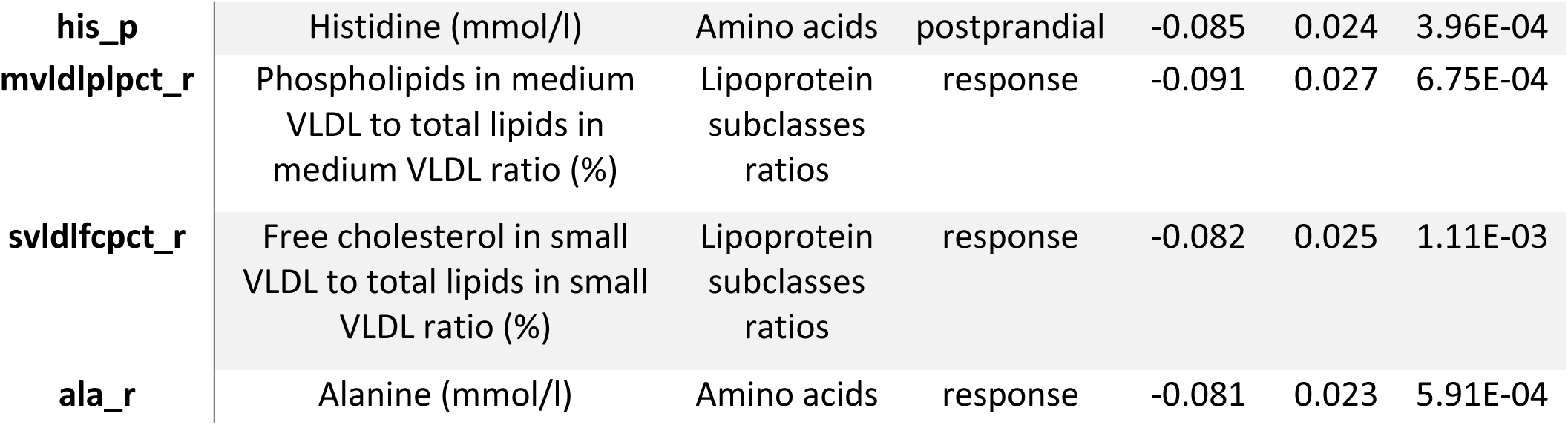
Metabolite traits causally associated with BMI. MR association summary statistics for the 20 metabolite traits associated with BMI. Provided are the trait name, metabolite description and units, metabolite class as defined by Nightingale Heath, the dietary state, and the effect estimate, standard error, and P-value. The effect estimates are in normalized standard deviation units of change per 1 unit change of BMI (kg/m^2^).

#### MR results - fasting

A total of 87 fasting traits exhibited nominal evidence of an association with BMI in the general population, five remained after correction for multiple testing (**Table 2**, **Figure 6, S3 File**). The strongest association observed was with the ratio of linoleic acid to total fatty acids (lafa; beta = -0.106, se = 0.025, P = 1.80×10^-5^). This was followed closely by 4 other inverse associations: the ratio polyunsaturated fatty acids (pufafa; beta = -0.081, se = 0.024, P = 6.80×10^-4^) to total fatty acids, the ratio omega-6 to total fatty acids (faw6fa; beta = -0.081, se = 0.024, P = 6.97×10^-4^) and the abundance of citrate (cit; beta = -0.079, se = 0.024, P = 8.63×10^-4^). In addition, one fasting metabolite showed evidence of a positive association with BMI, the branched-chain amino acid leucine (leu; beta = 0.068, se = 0.020, P = 5.62×10^-4^). If data is summarized by metabolite annotation class (**Table S1** in **S1 File**), we find that the classes with the largest average absolute BMI effect are ketone bodies (average beta = -0.047, n = 2), amino acids (0.036, n = 8), inflammation (0.029, n = 1), glycerides and phospholipids (0.027, n = 9) and lipoprotein subclasses (0.023, n = 98). Those with the smallest average absolute effect are lipoprotein particle size (0.009, n = 3), lipoprotein subclass ratios (0.003, n = 70), and cholesterol (0.002, n = 9) (**S10 Fig.**).

#### MR results - postprandial

A total of 96 postprandial metabolites exhibited nominal evidence of an association with BMI, 12 remained after correcting for multiple testing (**Table 2**, **Figure 6, S3 File**). Like the fasting metabolite traits the fatty acid ratios linoleic acid (lafa; beta = -0.092, se = 0.025, P = 2.0×10^-4^), omega-6 (faw6fa; beta = -0.080, se = 0.024, P = 7.83×10^-4^), polyunsaturated fatty acids (pufafa; beta = -0.077, se = 0.024, P = 1.10×10^-3^) and citrate (cit; beta = -0.105, se = 0.022, P = 2.83×10^-6^), who had the largest effect, each also had an inverse relationship with BMI (**Figure 5-6**). In addition, the amino acid histidine (his), large HDL cholesterol (l.hdl.c) and free cholesterol (l.hdl.fc) decreased with increases in BMI. Further, the ratio of cholesterol (l.hdl.c.pct), cholesterol esters (l.hdl.ce.pct), and free cholesterol (l.hdl.fc.pct) to total lipids in HDL decreased with increases in BMI. Finally, the ratio of free cholesterol to total lipids in IDL (idl.fc.pct) and the ratio of phospholipids to total lipids in medium VLDL (m.vldl.pl.pct) decreased with increases in BMI. If data is summarized by metabolite annotation class (**Table S1** in **S1 File**), we find that the classes with the largest average absolute BMI effect are glycolysis related metabolites (average beta = -0.046, n = 3), inflammation (0.40, n = 1) and lipoprotein subclasses (0.02, n = 98). The average effect among amino acids is not negative (- 0.019, n = 8) in opposition to that seen in fasting data (**S10 Fig.**).

#### MR results - response

A total of 18 response metabolites exhibited nominal evidence of an association with BMI, three remained after correcting for multiple testing (**Table 2**, **Figure 6, S3 File**). The largest effect was observed for the amino acid alanine (ala: beta = -0.081, se = 0.023, P = 5.91×10^-04^). Data suggested that increases in BMI attenuates the increase in abundance of the amino acid alanine after a liquid mixed meal (**Figure 5-6**) – consistent with observational analyses above (**Figure 4**). The other two response traits that associated with BMI are the ratio of phospholipids in medium VLDL to total lipids in medium VLDL (m.vldl.pl.pct; beta = 0.091, se = 0.027, P = 6.75×10^-4^) and the ratio of free cholesterol in small VLDL to total lipids in small VLDL (s.vldl.fc.pct; beta = 0.082, se = 0.025, P = 1.11×10^-3^). As a class, amino acids (average beta = -0.059, n=8) had the largest average BMI response effect estimate (**S10 Fig.**). This was followed by the ketone bodies acetate and beta-hydroxybutyrate (mean beta = 0.048, n = 2) and the glycolysis related metabolites citrate, glucose, and lactate (mean beta = -0.042, n = 3).

### Sub-population and sensitivity analyses

All observational and MR analyses were repeated in each of the two sub-populations and in the NEO cohort without the inclusion of weights. These analyses allowed us to evaluate the variability in effect estimates measured in a random (Leiderdorp) and biased (Leiden) population sample as well as the effectiveness of the weights in our primary analyses (wNEO). We found that effect estimates from the Leiderdorp and the wNEO frameworks are strongly correlated (observational Pearson’s r = 0.983, MR = 0.855) and correlation coefficients weakened when compared to the un-weighted NEO and the Leiden frameworks (**S11 Figure**). Decreases in agreement between analyses as the sample population mean BMI shifts would indicate that there are either un-accounted confounders influencing the results or that the relationship between BMI and metabolite trait variation is not always linear.

To evaluate the influence of confounders on MR effect estimates we reran the association analysis in the wNEO data set. Three additional covariates were included in the model (smoking, on a weight loss diet, and PC3) and 101 sample with a peroxidation sample quality flag, a sample quality metric identified by Metabolon, were removed. Overall primary (wNEO) observational (Pearson’s r = 0.998) and MR (Pearson’s r = 0.963) effect estimates correlate strongly with those from the sensitivity analysis (**S7 File**). A more comprehensive description of these analyses is in **S1 Text**.

## Discussion

This study provides effect estimates for observational and MR associations between BMI and metabolites in a (1) fasted, (2) postprandial, and (3) response state, using individual level data from a middle-aged cohort of 5517 individuals of Northern European ancestry including a liquid mixed meal challenge. Each dietary state provides a unique examination at metabolite variation, the value of which are actively being explored. Broad observational associations were observed between BMI and metabolites (69% of tested traits). In addition, MR estimates were largely concordant with the observational estimates (**Figure 3D**). This is consistent with previous research and suggests that, at least for this lipidomic platform, observational estimates are reasonably well aligned to MR effect estimates (18). This generalization holds less true for response traits than for postprandial and fasting trait (**Figure 3D**). After correcting for multiple testing, 20 metabolite traits maintain evidence of a causal BMI effect (**Figure 6**). This includes the inverse association with fasting and postprandial ratios of linoleic acid (lafa), omega-6 (faw6fa), and polyunsaturated fatty acids (pufafa) to total fatty acids and fasting and postprandial citrate (cit) abundance. In addition, the amino acid leucine has a positive MR association with BMI in the fasting state, histidine has an inverse MR association with BMI in the postprandial state and alanine has an inverse MR association with BMI in the response state.

We compared our observational effect estimates to fifty-seven matching fasting metabolites measured using the same platform, in a young adult cohort (18). Overall effect estimates correlated well (Pearson’s r = 0.82), with the caveat that estimates cannot be directly compared given differences in data transformations between the studies. However, there are 37 shared associations, 6 unique to this study, 12 unique to the other and 2 yielding no association in either study (**Figure 3B**). Five of the shared associations were different across studies (total cholesterol, sphingomyelins, phosphatidylcholines, phosphoglycerides, and linoleic acid). Among the 12 metabolites unique to existing work before that presented here, are 10 metabolites that previously exhibited a strong positive effect but showed no evidence for an association or negative point estimates here (total lipids in IDL, large LDL, medium LDL and small LDL, total cholesterol in IDL and LDL, and four fatty acid metabolites omega-3, omega-6, polyunsaturated fatty acids, and docosahexaenoic acid). These differences may be the product of non-linear associations between these metabolites and BMI in combination with age effects and differences in the distribution of BMI between these studies. Despite these difference, 57 fasting MR estimates that did overlap with previous work showed strong correlation between the two studies (Person’s r = 0.862, P = 7.99×10^-18^; **S12 Fig.**), with one shared fasting MR association – the amino acid leucine.

Given the prevalence of lipoproteins and their lipids on this platform we looked for the expected atherogenic lipoprotein profile – that is decreases in HDL and increases in non-HDL lipoproteins with increase in BMI (63–66). Both observational and MR associations between BMI and fasting and postprandial dietary states do suggest a worsening atherogenic lipoprotein profile with BMI (**Figure 7, S13 Fig,** and **S14 Fig.**). As illustrated with the fasting observational effect estimates in **Figure 7**, the atherogenic VLDL lipoproteins are higher with increases in BMI while non-atherogenic lipoproteins – or those involved in the reverse cholesterol transport system (HDL), were lower. In contrast to expectations, other atherogenic lipoproteins IDL and LDL, largely do not associate with BMI. In addition, triglycerides in non-atherogenic medium HDL and small HDL increase with increases in BMI. Further, as HDL lipoprotein density increases the inverse relationship with BMI weakens and as seen with small HDL the association becomes positive (**Figure 7**). This is consistent with the observation above that the strongest inverse association with BMI is that with average diameter of HDL particles, suggesting that as BMI increases there is a associated shift toward smaller HDL particle size. Along with these changes, other observations were consistent with obesity influencing lipid profiles that are associated with cardiometabolic disease (17,25,67). These included increases in apolipoprotein B, decreases in apolipoprotein A-1 and increases in the ratio of B to A-1 with increases in BMI, in both the fasting and postprandial states (65,68).

**Figure 7:**
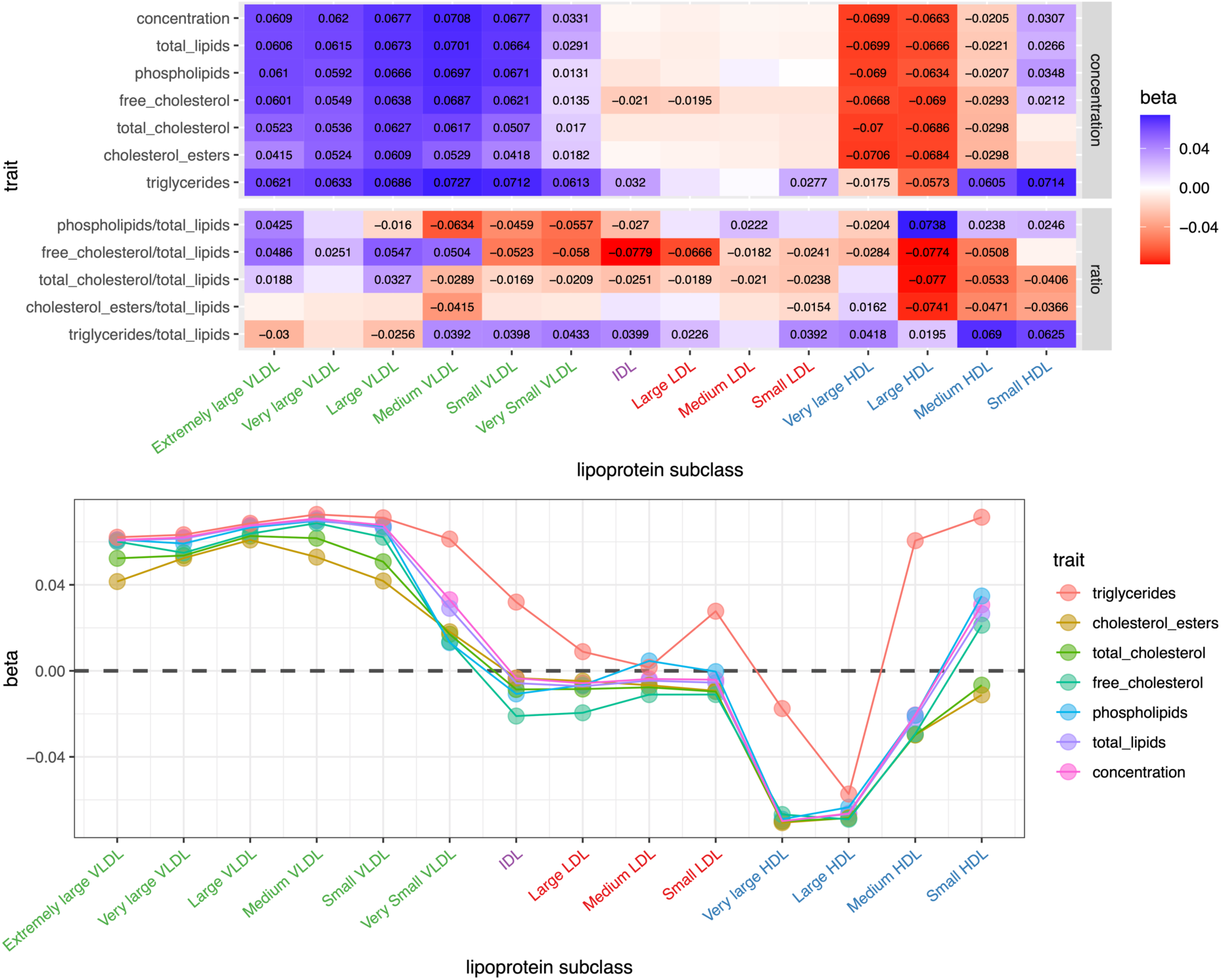
Fasting lipoprotein cross-sectional profile. (Upper) A tile plot of cross-sectional effect estimates for lipoproteins and lipoprotein ratios in the fasting state. Tiles with an effect estimate provided in text are those associated with BMI (p<1.16×10^-3^). The lipoproteins are organized by size or density along the x-axis, and the component or ratio being measured is along the y-axis. (Lower) A dot plot or profile of cross-sectional effect estimates for lipoproteins (x-axis) ordered by lipoprotein size or density with effect estimates (y-axis). The component or measurement of each lipoprotein are defined by the color as described in the key.

The MR association of BMI across branched chain (BCAA) and aromatic (AAA) amino acids were similar with positive effects in the fasting state, no effect in the postprandial state, and an inverse association in the response state (**Figure 2 and 5**). Meanwhile the dietary profile of (glucogenic) amino acids suggest no effect in the fasting state and inverse effects in both the postprandial and response states. These observations may suggest that BMI may have a causal influence on the synthesis or metabolism of amino acids in general, particularly regarding their inverse association with BMI in the response state (**Figure 2 and 5**). BCAAs and AAAs have long been associated with obesity, glucose, insulin (resistance), and type 2 diabetes (69–75). In support of early observations, numerous recent studies that have shown associations between amino acids and BMI, visceral adipose tissue and weight change (18,36,76–78), between BCAAs and AAAs with insulin resistance and type 2 diabetes (79–82), used MR to show that insulin resistance increase BCAAs concentration (83) and used a randomized control trail to show that restricting BCAAs can improve glucose tolerance and reduce fat accumulation (84). Indeed evidence now suggests a potential causal pathway from BMI to type 2 diabetes via the intermediate traits of insulin resistance and BCAAs, respectively (83). Observations here reinforce the causal association between BMI and amino acids, with the additional observations that postprandial abundance of the glucogenic amino acids decreases with BMI and that the relative change in amino acid abundance in response to a liquid mixed meal decreases with increases in BMI.

As the study was centered in the Netherlands, study samples were limited to individuals of Northern European ancestry and as such inferences made from results should be limited to populations of similar ancestry and environment. Sampling was limited to individuals of middle age (range 44 to 66 years and a mean of 56, Table 1). Whilst this provides specificity to a middle age population it also limits the results to this population age range as well. The NEO population sampling was performed in two distinct batches, one focused on oversampling individuals of high BMI (Leiden) and a second a random and representative sampling of the population (Leiderdorp). The product of this led to the use of weighted linear regression analyses to maintain sample size and power, while also providing estimates that are representative of the study population at large. The validity of this assumption rests in the credibility of the weights, which were specifically designed to make the BMI distribution of the Leiden sample mirror that of the Leiderdorp sample (85). We have illustrated through comparison with the Leiderdorp sample that effect estimates are consistent across the two analyses, imparting support to the weighted analysis (**S11 Figure)**. With these factors, there was just a single postprandial time point evaluated here limiting the inferences that can be drawn. It is reasonable to speculate that an earlier time point, or a complete time series would provide additional information about how BMI might influence postprandial variation. Finally, while a single complex meal standardizes the complexities in evaluating metabolite response and postprandial abundances alternative meals could provide novel insights in the interplay between BMI, nutrition, and metabolite variation.

In conclusion, using a middle-aged cohort of 5517 individuals we have demonstrated that effect estimates for BMI on metabolite traits in the fasting, postprandial and response dietary states. We have been able to show that results are broadly correlated between observational and one-sample Mendelian randomization analyses and in the context of the numerous documented associations between BMI, metabolites and disease, this gives support to a conclusion that metabolites may act as intermediates – or at the least biomarkers of underlying driver physiology – between adiposity accumulation and disease. Furthermore, this work suggests that the dynamic metabolome may potentially flag common biological events which will systematically vary by BMI, which may be linked to the aetiology of disease and which are linked to life course events such as feeding and metabolic response.

## Data Availability

The individual-level data are not publicly available but access may be granted to researcher through an application procedure by contacting the scientific director of the NEO study, Renee de Mutsert.

https://www.lumc.nl/over-het-lumc/afdelingen/klinische-epidemiologie/obesity-related-diseases/

## Acknowledgements

The authors thank all individuals who participated in the Netherlands Epidemiology of Obesity study, and all participating general practitioners for inviting eligible participants. We also thank the NEO study group, Pat van Beelen and all research nurses for data collection, Petra Noordijk and her team for laboratory management, and Ingeborg de Jonge for data management of the NEO study. We acknowledge the support from the Netherlands Cardiovascular Research Initiative: an initiative with support of the Dutch Heart Foundation (CVON2014-02 ENERGISE). We thank Nutricia Research, Utrecht, The Netherlands, for providing the liqud mixed meal. Nicholas J. Timpson is a Wellcome Trust Investigator (202802/Z/16/Z), is the PI of the Avon Longitudinal Study of Parents and Children (MRC & WT 102215/2/13/2), is supported by the University of Bristol NIHR Biomedical Research Centre (BRC-1215-20011), the MRC Integrative Epidemiology Unit (MC_UU_12013/3) and works within the CRUK Integrative Cancer Epidemiology Programme (C18281/A19169). David A. Hughes is supported by N.J.Timpson’s Wellcome Investigator Award (202802/Z/16/Z), and along with Caroline Bull is also supported by the University of Bristol and UK Medical Research Council (MC_UU_00011/1 and MC_UU_00011/6). Dennis Mook-Kanamori is supported by Dutch Science Organization (ZonMW-VENI Grant 916.14.023)

