## Supplementary material for "The association between body mass index and metabolite response to a liquid mixed meal challenge": S1_Log.txt

I. Setting up your pipeline
	-Your data directory is: /Volumes/MRC-IEU-research/projects/wt1/wp1/010/working/data/source/release2/metaboprep/
	- Your metabolite data file is: source_metabolites_extracted.csv
		Your metabolite data file was identified as a previously processed flat text file.
	- Your provided feature annotation file to process is: source_metabolites_feature_data.txt
	- Your provided sample | batch annotation file to process is: source_metabolite_batch_data.txt
	- Your declared platform is: Other
	- Feature filtering: Your declared feature missingness level is: 0.2
	- Sample filtering: Your declared sample missingness level is: 0.2
	- Sample filtering: Your declared total peak area filter level, in standard deviations from the mean is: 5
	- Sample filtering: Your declared principal component (PC1 and PC2) exclusion, in standard deviations from the mean is: 5
	- Metabolite independence: Your declared tree cut height is: 0.5

	- Reading in your csv metabolite file
	- Reading in you txt feature annotation file
	- Reading in you txt sample annotation file
		- Assuming sample IDs are in column 1 and redefining rownames
III. Your data has been read in.

	-Your data has 5744 individuals and 606 metabolites.

	-There are also 3 sample annotation|batch variables.

	-There are also 10 feature annotation|batch variables.

IV. Estimating Summary Statistics On Raw Data Set.
	a. Estimating summary statistics for samples
		- Writing sample summary statistics to file.
	b. Estimating summary statistics for features.
		- Estimating the number of indpendent features.
			- 12 features excluded from analysis for (>20% missingness) n smaller than 4595.
			- Generating Correlation Matrix.
			- Generating Distance Matrix.
			- Constructing dendrogram.
			- Performing tree cut. Cut height defined at 0.5
			- Identifying independent features.
		- Writing feature summary statistics to file.
	c. Performing principle component analysis and identifying outliers.
		- Re-Writing sample summary statistics to file to include PCs.
		- Writing PC statistics to file.

V. Performing data filtering.
	a. Performing data filtering.
		- QCstep: estimate initial sample missingness.
		- QCstep: exclude those sample with missingness >= 80%.
			* 9 sample(s) excluded for extreme missingness.
		- QCstep: estimate initial feature missingness.
		- QCstep: exclude those features with missingness >= 80%.
			* 0 features excluded for extreme missingness.
		- QCstep: RE-estimate sample missingness.
		- QCstep: exclude those sample with missingness >= 20%.
			* 217 sample(s) excluded for user defined missingness.
		- QCstep: RE-estimate feature missingness.
		- QCstep: exclude those features with missingness >= 20%.
			* 3 feature(s) excluded for user defined missingness.
		- QCstep: estimate total peak area.
		- QCstep: exclude those features with TPA >= 5SD from the mean.
			* 0 samples excluded for user defined TPA SD from the mean.
		- QCstep: re-identify independent features through correlation analysis and dendrogram clustering.
			- using currently QCd data.
		- Estimating the number of indpendent features.
			- Generating Correlation Matrix.
			- Generating Distance Matrix.
			- Constructing dendrogram.
			- Performing tree cut. Cut height defined at 0.5
			- Identifying independent features.
			* 78 independent features identified.
		- QCstep: Perform Principle Componenet Analysis of currently QC'd data.
		- QCstep: Identify PC 1-2 outliers >= +/-5SD of the mean.
			* 0 samples excluded as PC outliers.
	b. Writing QC data to file.

VI. Estimating Summary Statistics on Filtered Data Set.
	a. Estimating summary statistics for Filtered samples
	b. Writing filtered sample summary statistics to file.
	c. Estimating summary statistics for filtered features.
		- Estimating the number of indpendent features.
			- Generating Correlation Matrix.
			- Generating Distance Matrix.
			- Constructing dendrogram.
			- Performing tree cut. Cut height defined at 0.5
			- Identifying independent features.
			- A total of 78 independent features were identified in the total filtered data set.
	d. Writing feature summary statistics to file.
	e. Performing principle component analysis on final filtered data set.
		 The number of informative principle components:
			 1. Cattel's Scree Test : acceleration factor = 2
			 1. Parrallel Analysis = 18
	f. Re-Writing filtered sample summary statistics to file to include PCs.
	g. Writing PC statistics to file.

VII. Generate Data Description pdf report.
