## Supplementary material for "The association between body mass index and metabolite response to a liquid mixed meal challenge": S1_Text.pdf

### Supplementary Methods and Results

**Manuscript**
The relationship between body mass index and metabolite response to a standardized meal challenge.

**Authors**
David A. Hughes[1,2*], Ruifang Li-Gao[3], Caroline J. Bull[1,2], Renée de Mutsert[3], Frits R. Rosendaal[3], Dennis O. Mook-Kanamori[3], Ko Willems van Dijk[3], Nicholas J. Timpson[1,2.]

**Affiliations**
[1] MRC Integrative Epidemiology Unit at the University of Bristol, UK
[2] Population Health Science, Bristol Medical School, University of Bristol, UK
[3]Department of Clinical Epidemiology, Leiden University Medical Center, Leiden, the Netherlands
[4]Department of Public Health and Primary Care, Leiden University Medical Center, Leiden, the Netherlands
[5]Department of Human Genetics, Leiden University Medical Center, Leiden, the Netherlands
[6]Department of Internal Medicine, Division of Endocrinology, Leiden University Medical Center, Leiden, the Netherlands

### Supplementary Methods

#### Metabolite data quality control

The initial NEO metabolite data set contained data for 5,744 individuals, 229 fasting metabolite, 229 postprandial metabolites, and 148 previously derived (38) ornls response metabolite traits. We used the R package metaboprep to perform quality control on the data prior to analysis. The following parameter values were used when running metaboprep: (1) feature missingness 0.2, (2) sample missingness 0.2, (3) total peak area standard deviation 5, (4) principal component (PC) outlier standard deviations 5, (5) tree cut height 0.5, (6) derived variable exclusion FALSE. Metaboprep performs an initial exclusion of samples with missingness greater than or equal to 80% (n = 9), followed by exclusion of features with missingness greater than equal to 80% (n = 0). After this initial filtering, the data set is filtered using the parameters defined above. This will filter (1) samples (n = 217) and then (2) features (n = 3) with greater than or equal to 20% missingness, (3) then filter samples with a total sum abundance (TSA) at complete features (no missingness) that fall beyond 5 standard deviations from the mean of the observed TSA distribution (n = 0), and then (4) filter samples (n = 0) that are 5 standard deviations from the mean of the first 'i' informative PCs, here defined by the Cattel's Scree test acceleration factor (i = 2). Parameter 5 defines the clustering dendrogram tree cut height which was used to estimate the number of clusters, and then used to identify the representative or principal variables in the data set. These principal variables were used in the data filter PC analysis used by parameter 4 above. Derived variables or features such as ratios that are derived from multiple features in the data set were (6) not excluded from any analyses. Given the defined *metaboprep* parameters 226 samples and 3 metabolites, all ornls response traits, were filtered from the data set. The log file (**S1 Log**) and report (**S1 Report**) generated by *metaboprep* are available in supplementary data.

Following metaboprep quality control, additional quality control steps were taken. First, all zero values were turned into NAs. Second, for each metabolite (in the fasting and postprandial state, individually) any sample with a value 10 interquartile distances from the median was turned into NAs. This QC-step removes all extreme observations that are unrealistic given the empirical distribution of a single metabolite trait. Third, for each metabolite we used the expectation that fasting and postprandial data are correlated to estimate a delta value (postprandial - fasting), from which all samples that fell five interquartile distances from the median of that metabolite's delta distribution were turned into NAs - in both the fasting and postprandial data. This QC-step removes all extreme observations that are unrealistic given the observed, empirical, correlated nature of this bivariate data. See **Fig. A** for an example and see **S2 File** for pre- and post-QC scatter plots for all metabolites.

#### Figure A: Raw and QC'd fasting and postprandial scatter plots for S-HDL-C

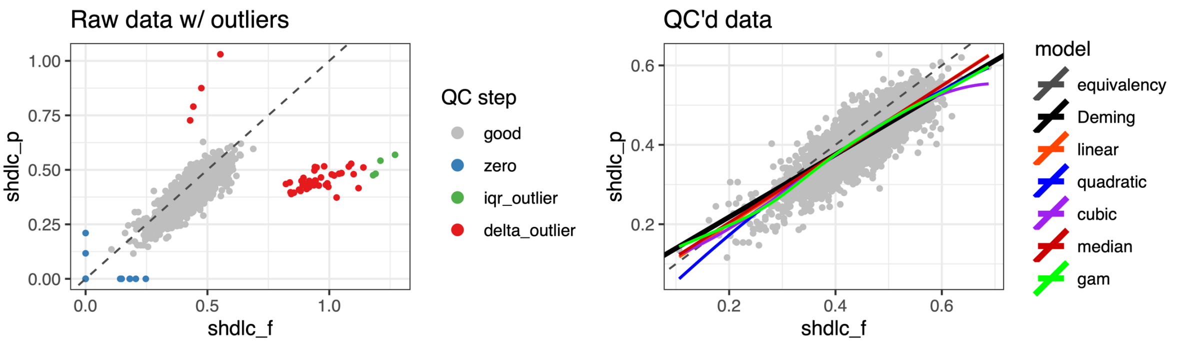

**Figure A: Raw and QC'd fasting and postprandial scatter plots for S-HDL-C.** (***Left***) A scatter plot for fasting (x-axis) and postprandaial (y-axis) cholesterol in small HDL (S-HDL-C). An equivalency line (x = y) is illustrated as a dashed grey line. Each dot is a sample, those colored blue had a zero value and were turned into NA, those colored green were identified as outliers (10 interquartile distances from the median) in one of the two dietary states and turned into NA in that dietary state, those colored red were identified as delta outliers (5 interquartile distances from the median). Delta was estimates as the postprandial value minus the fasting value. Those samples colored as grey were not altered. (***Right***) is a scatter plot for fasting (x-axis) and postprandaial (y-axis) total lipids in small HDL (S-HDL-L) after quality control. An an equivalency line (x = y) is illustrated as a dashed grey line. In addition, six linear models were fit to the data and are plotted. They are a (black) a Deming regression, (orange) a linear model, (blue) a linear model were the independent term or fasting data was fit as a quadratic term, (purple) a linear model where the fasting data was fit as a cubic term, (red) a median regression, and (green) a generalized additive model or GAM where the fasting data was fit as a smooth. Each of these models were fit to evaluate how much variability was present among each model type. A plot like the above is available for each metabolite in the study in **S2 File**.

#### Effective number of tested metabolites

There are 687 metabolite traits in the data set (fasting = 229, postprandial = 229, response = 229) but these traits are not strictly independent. First, given the presence of both fasting and postprandial data, and a response trait derived from the two, we would expect there to be a degree of dependency. Second, given the focus on lipids and lipoproteins on this platform we also expect an abundance of inter-correlated structure (**S3 Fig.**). As such we used the R package *iPVs* (https://github.com/hughesevoanth/iPVs) to estimate the effective number of metabolites in the data set. The effective number represents an estimate of the number of representative or independent traits present in the data (47–49). The method implemented by iPVs provides an estimate by the iterative construction of a hierarchical clustering dendrogram followed by a tree cut to identify clusters of correlated variables and identification of a principal variable (PVs) for each cluster. The iPVs() function parameters were set to: "spearman" for the correlation matrix, "R" for the distance matrix, "complete" for the hierarchical cluster method, and 0.5 for the tree cut height. The distance matrix produced is equal to one minus the absolute Spearman's rho. At a tree cut height of 0.5, metabolites with a Spearman's rho greater than 0.5 cluster together. If the tree cut height was, for example, set to a value of 0.8, then metabolites with a Spearman's rho greater than 0.2 would cluster together. In total 43 clusters or representative variables in the NEO metabolite data set were identified. Resultantly our data reduced, study-wide Bonferroni (BF) corrected p-value was set to 0.05/43 or $1.163 \times 10^{-3}$.

##### Identification of possible confounders

To identify possible confounders, or variables that may violate MR IV assumption number two (independence) we tested, in a univariate fashion, for an association between (a) sampling date and BMI and BMI-PGS, (b) study covariables and BMI, (c) study covariables and BMI-PGS and (d) study covariables and metabolite traits (Table S4 in **S1 File**). An ANOVA analysis of the univariate linear models was then used to partition the sums of squares and estimate the variance explained, in the form of an eta-squared statistic ($\eta^2$). Eta-squared ($\eta^2$) is a measure of the variance explained and can be derived from sums of squares or deviances extracted from ANOVAs. In addition, an ANOVA F-test was used to estimate a p-value for each association. These analyses were carried out for the NEO cohort as a whole and for each of the two NEO sub-samples and provides a means to identify possible confounders that may invalidate the MR framework.

#### Supplementary Results

##### Possible confounders

We performed a complete assessment of the correlation between BMI and BMI-PGS with all study covariables to identify those variables that could be possible confounders in association analyses. All covariables were chosen for inclusion because of a potential correlation with BMI and metabolite trait variation. Indeed, we observed that 60 of the tested 91 covariables are associated with BMI (wNEO, $P < 9.26 \times 10^{-4}$; Table S4 in **S1 File**; **S7 Fig.**). Those with the strongest association with BMI are the study sampling variables visit date and sub-population (already included as model covariates), followed by the obesity adjacent traits weight and hip and waist circumference, and then basal metabolic rate, glucose metabolism, resting energy expenditure (kcal/day), mean oxygen production (VO2, ml/min) and mean carbon dioxide production (VCO2, ml/min). In addition, smoking as measured by packyears associates with BMI, explaining 3.2% ($\eta^2$) of the variation in BMI (wNEO, ANOVA F-test $P = 1.28 \times 10^{-37}$), as does education (edu_level, $\eta^2 = 0.044$, $P = 1.96x10^{-47}$), and household income (income_hh, $\eta^2 = 0.015$, $P = 1.52x10^{-14}$, **Table S4** in **S1 File**).

In contrast, the MR instrumental variable (BMI-PGS) is associated with 17 covariables (wNEO, $P < 9.26x10^{-4}$, **S8 Fig.**), five of which are obesity adjacent traits, three are sampling and sub-population structure variables, three smoking variables, 4 diet variables, a calorimetry variable, and a sample quality variable. They are, in order of association strength: hip circumference ($\eta^2 = 0.039$, $P = 2.05x10^{-49}$), weight ($\eta^2 = 0.038$, $P = 9.97x10^{-49}$), waist circumference ($\eta^2 = 0.035$, $P = 1.76x10^{-44}$), sub-population ($\eta^2 = 0.020$, $P = 3.61x10^{-26}$), basal metabolic rate ($\eta^2 = 0.016$, $P = 1.00x10^{-20}$), waist-to-hip ratio ($\eta^2 = 0.011$, $P = 7.43x10^{-15}$), visit date ($\eta^2 = 0.029$, $P = 1.56x10^{-14}$), principal component 3 ($\eta^2 = 0.006$, $P = 5.03x10^{-09}$), smoking packyears ($\eta^2 = 0.006$, $P = 4.16x10^{-08}$), on a weight loss diet ($\eta^2 = 0.004$, $P = 6.40x10^{-07}$), imputed smoking packyears ($\eta^2 = 0.005$, $P = 7.63x10^{-07}$), type of diet ($\eta^2 = 0.007$, $P = 1.24x10^{-06}$), on a diet last month (yes|sometimes|no, $\eta^2 = 0.004$, $P = 8.04x10^{-06}$), smoker (never|former|current, $\eta^2 = 0.004$, $P = 1.01x10^{-05}$), food rule description ($\eta^2 = 0.037$, $P = 9.60x10^{-05}$), mean $CO_2$ production ($\eta^2 = 0.010$, $P = 6.04x10^{-04}$), and the sample quality flag for signs of peroxidation ($\eta^2 = 0.002$, $P = 8.30x10^{-04}$, **Table S4** in **S1 File**). The association between BMI-PGS and principal component three is partially driven by its association with sub-population structure ($\eta^2 = 0.032$, $P = 1.71x10^{-41}$, **Fig. B**). All 17 covariables associated the BMI-PGS were also tested for an association with each metabolite trait in a univariable linear model. Results illustrated broad association across all traits and covariables, defining each covariable as a confounder in MR analysis (**S9 Fig.**).

#### Figure B: Sub-population association with PC3

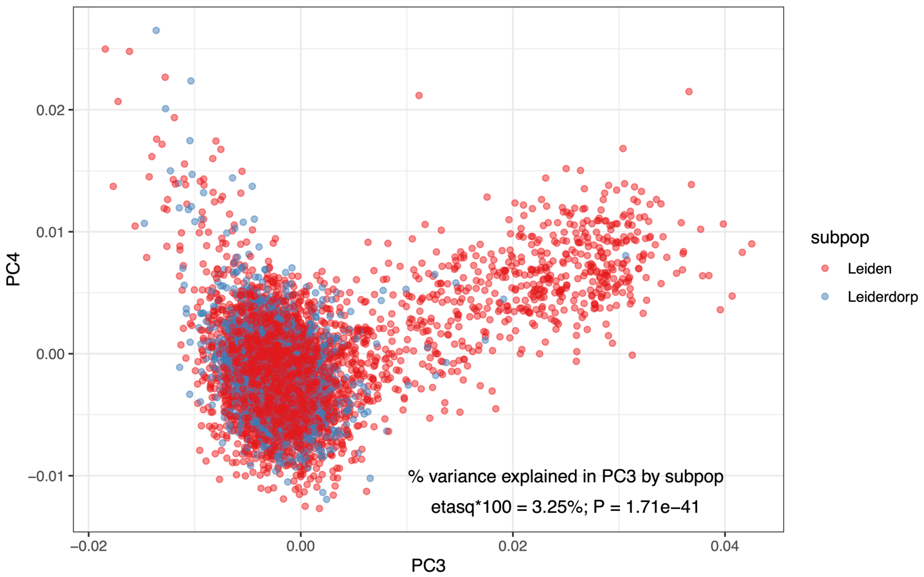

**Figure B: Sub-population association with PC3.** A scatter plot of NEO study samples on principal component three and four, as provided as covariables by NEO data managers. Each dot is an individual that is colored by the sample sub-population Leiden (red) and Leiderdorp (blue). PCs one ($\eta^2 = 0.002$, P = $9.33x10^{-04}$), three ($\eta^2 = 0.032$, P = $1.71x10^{-41}$), and four ($\eta^2 = 0.006$, P = $3.16x10^{-08}$) are each correlated with sub-population, but PC3 has the strongest association that is also associated with the MR instrumental variable BMI-PGS making it a possible confounder in MR analysis, but only if sub-population was not already included in the MR models as a covariate.

#### Sub-population analyses

All observational and MR analyses were repeated in each of the two sub-populations and in the NEO cohort without the inclusion of weights. The (i) randomly sampled Leiderdorp sub-population (n=1406) provides a means to verify the point estimates derived from the primary weighted NEO (wNEO) results presented above. The (ii) unweighted NEO cohort provides an evaluation of the weights, and the (iii) Leiden sub-population sample (n=4111) provides an evaluation of effect estimates derived from a sampling biased for elevated BMI.

First, observational effect estimates from the wNEO analysis strongly correlate with those from the Leiderdorp sub-sample, with a Pearson's r of 0.983. The correlation does reduce when compared to the un-weighted NEO analysis (r = 0.96) and the biased Leiden sample (r = 0.91, **S11 Fig.**). Second, MR effect estimates remain strongly correlated between the wNEO and Liederdorp sample (r = 0.855), but as the shift in sample population BMI increases the correlation reduces. When compared to the un-weighted NEO sample the Pearson's r is 0.445 (P = $1.10x10^{-34}$) and even becomes negative when compared to the Leiden sample (Pearson's r = -0.107, P = $4.84x10^{-03}$; **S11 Figure**). The comparison to the randomly sample Leiderdorp sample would suggest that the weights used in the wNEO analysis did not introduce any strong error in effect estimates. Yet, the decrease in congruency between analyses as the sample population mean BMI shifts would indicate that there are either un-accounted for confounders influencing the results or that the relationship between BMI and metabolite trait variation are not linear.

#### Sensitivity analyses

To evaluate the influence of confounders on MR effect estimates we reran the association analysis in the wNEO data set. We assumed that the association between BMI-PGS and hip circumference, weight, waist circumference, basal metabolic rate, waist-to-hip ratio, and mean $CO^2$ production were the product of biological similarities with our exposure of interest, BMI, and not confounders. We did include a smoking variable (packyears), a diet variable (on a weight loss diet), and PC3 as new covariates in the MR analysis. We also excluded the 101 samples with a peroxidation flag on their samples. Overall primary (wNEO) observational (Pearson's r = 0.998) and MR (Pearson's r = 0.963) effect estimates correlate strongly with those from the sensitivity analysis (**S3 File**). Further no effect estimates differ between the primary and sensitivity models (z-test, $P< 0.05$) indicating that the inclusion of the additional confounders had minimal effect on the association statistics (**Fig C** and **D**).

### Figure C: Forest plot of observational MR sensitivity analyses

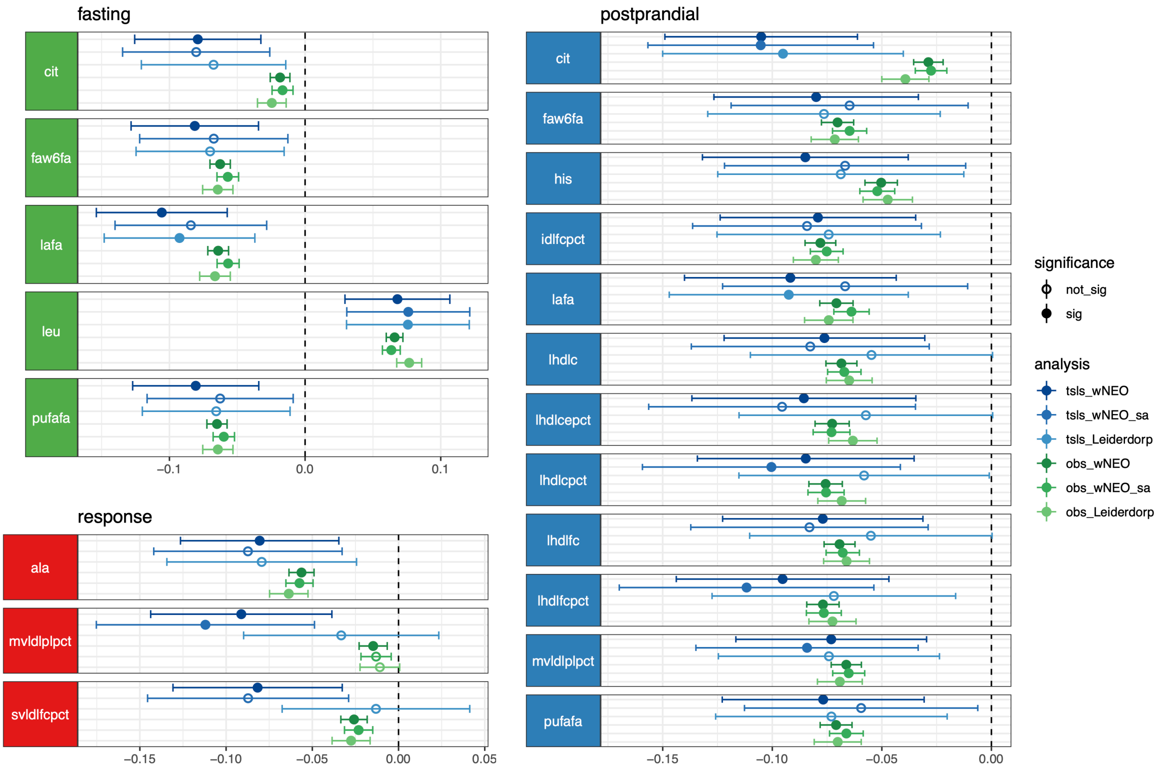

**Figure C: Forest plot of observational and MR sensitivity analyses.** A forest plot of effect estimates (points) and 95% confidence intervals (whiskers) for MR (tsls,blue) and observational (obs, green) effect estimates for the weighted NEO (wNEO), weighted NEO sensitivity analyses (wNEO_sa), and the Leiderdorp (sub-)populations.

#### Figure D: Correlation between wNEO and wNEO sensitivity analysis MR estimates

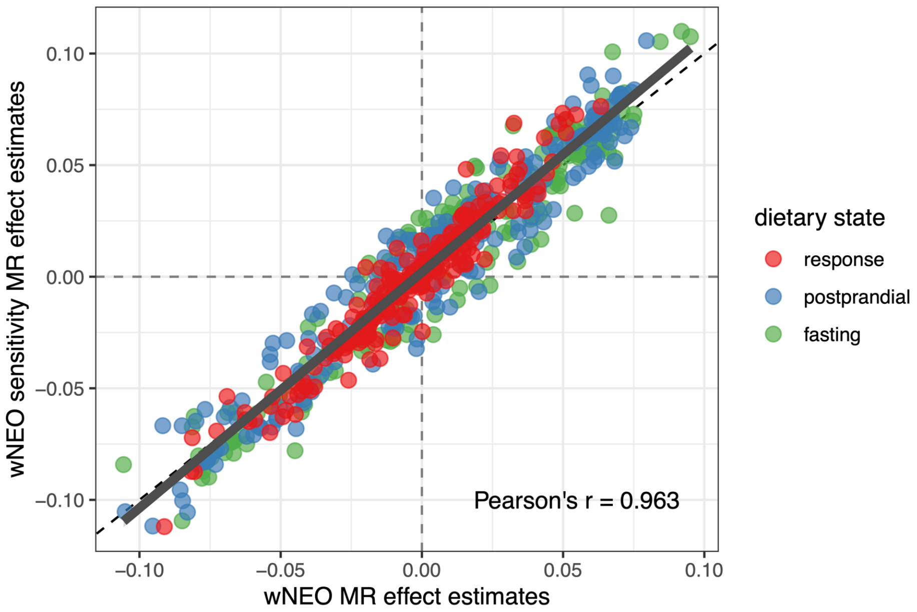

**Figure D: Correlation between wNEO and wNEO sensitivity analysis MR estimates.** Scatter plot of MR effect estimates in the weighted NEO (x-axis) and weighted NEO sensitivity analysis (y-axis). Point estimates for all metabolites in each of the three dietary states response (red), postprandial (blue), and fasting (green) are provided.
