## Supplementary material for "The association between body mass index and metabolite response to a liquid mixed meal challenge": Supplementary_Figures.pdf

**Manuscript**
The relationship between body mass index and metabolite response to a standardized meal challenge.

**Authors**
David A. Hughes[1,2*], Ruifang Li-Gao[3], Caroline J. Bull[1,2], Renée de Mutsert[3], Frits R. Rosendaal[3], Dennis O. Mook-Kanamori[3], Ko Willems van Dijk[3], Nicholas J. Timpson[1,2.]

**Affiliations**
[1] MRC Integrative Epidemiology Unit at the University of Bristol, UK
[2] Population Health Science, Bristol Medical School, University of Bristol, UK
[3]Department of Clinical Epidemiology, Leiden University Medical Center, Leiden, the Netherlands
[4]Department of Public Health and Primary Care, Leiden University Medical Center, Leiden, the Netherlands
[5]Department of Human Genetics, Leiden University Medical Center, Leiden, the Netherlands
[6]Department of Internal Medicine, Division of Endocrinology, Leiden University Medical Center, Leiden, the Netherlands

### Supplementary Figures

#### S1 Figure: Distribution of BMI

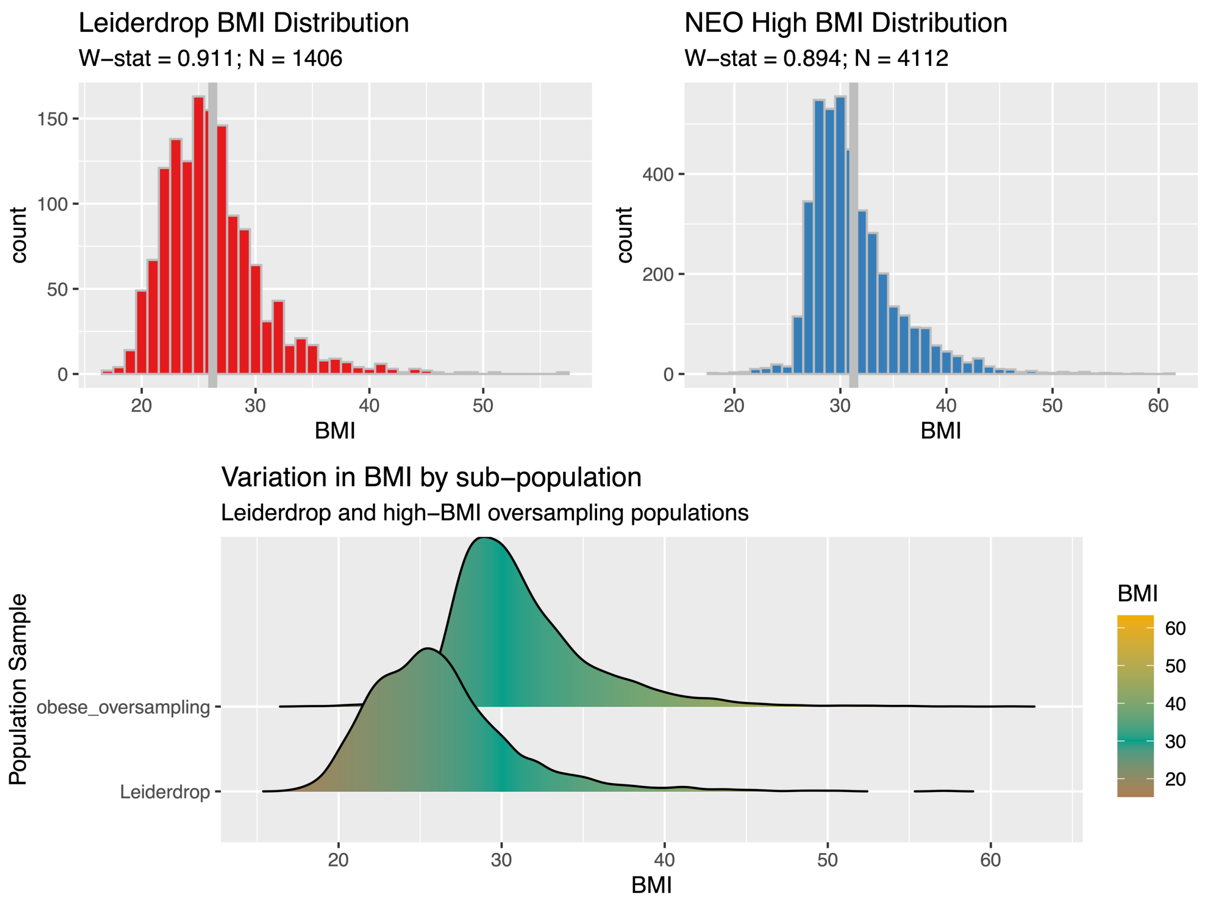

**S1 Figure: Distribution of BMI by sub-population.** The distribution of BMI in the Leiderdorp (red, top left) and the Leiden (blue, top right) subpopulations, and compared to each other (lower).

#### S2 Figure: BMI and PGS by visit data

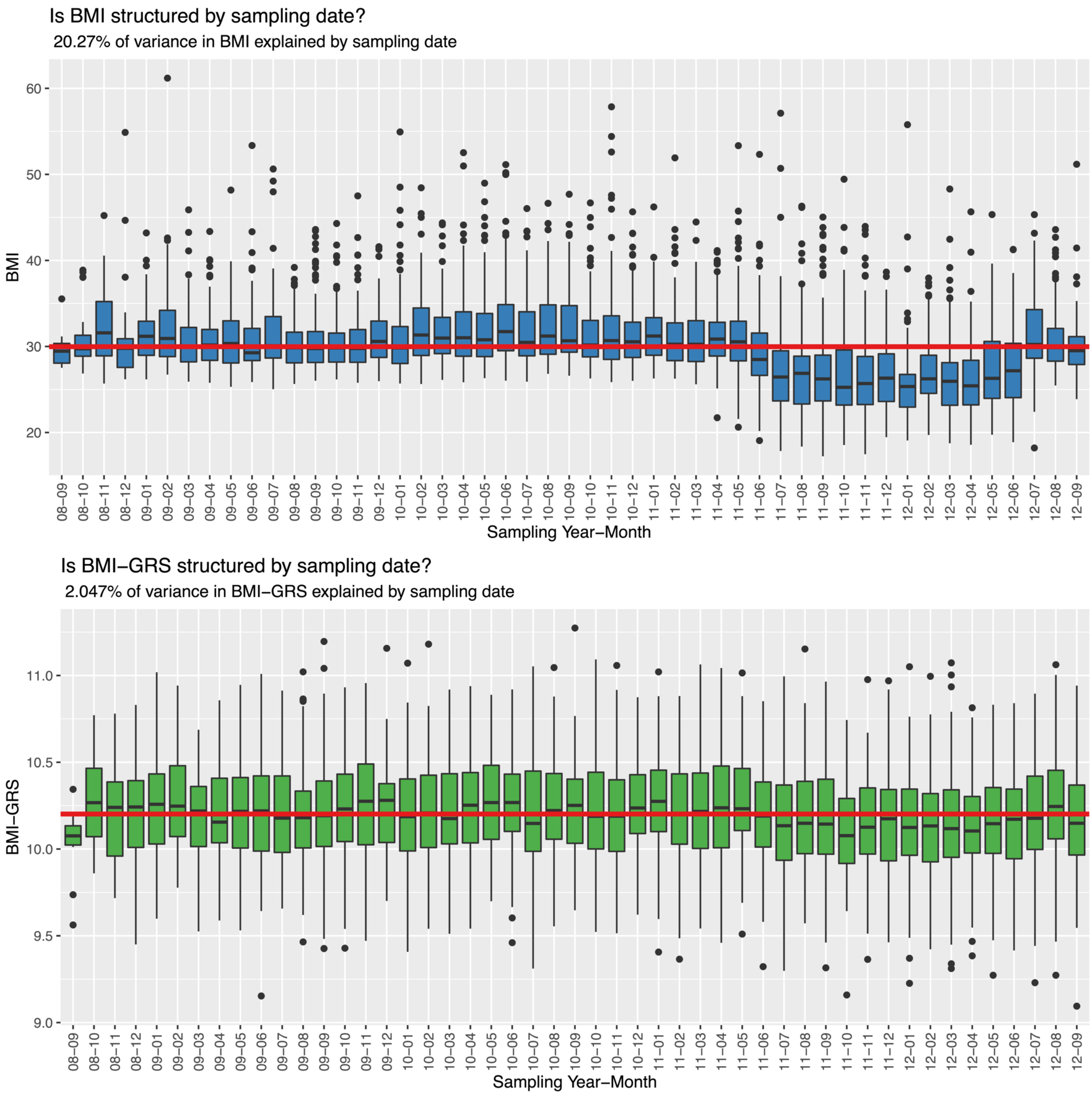

**S2 Figure: Box plots BMI distributions by sampling data.** NEO sampling dates (month-year) along the x-axis with the mean (box vertical line), 1st and 3rd quartile (box) and 1.5*IQR of box (whiskers) illustrated. The red vertical line illustrates the mean value of the total sample. Upper plot illustrates variation in BMI. The lower plot illustrates the variation in the instrumental variable – BMI-PGS. Defined in each sub-header is the proportion of variation in BMI (upper) or BMI-PGS (lower) explained by sample date.

#### S3 Figure: Metabolite and covariable clustering dendrogram.

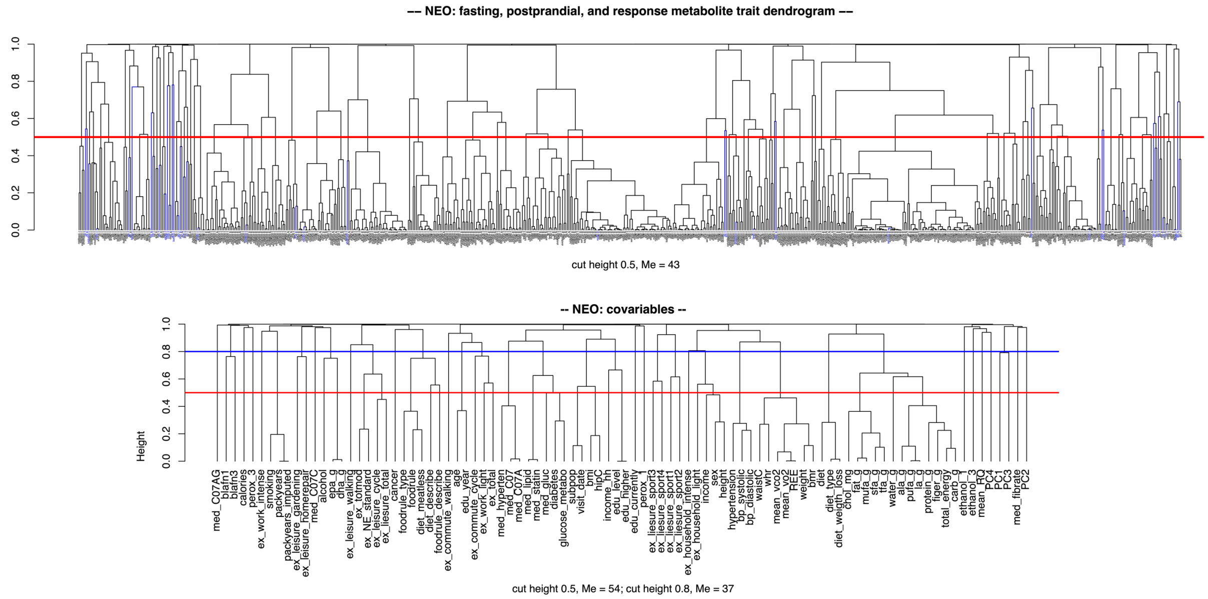

**S3 Figure: Metabolite and covariable clustering dendrogram.** Hierarchical clustering dendrogram of metabolites (top) and study covariables (bottom). Leafs colored blue indicate principal variables, as identified by the iPVs R package. The red horizontal line, at a height of 0.5, marks the tree cut height used to identify clusters by iPVs. The y-axis is equivalent to 1-absolute Spearman's rho.

#### S4 Figure: Mean lipid estimates across lipoproteins

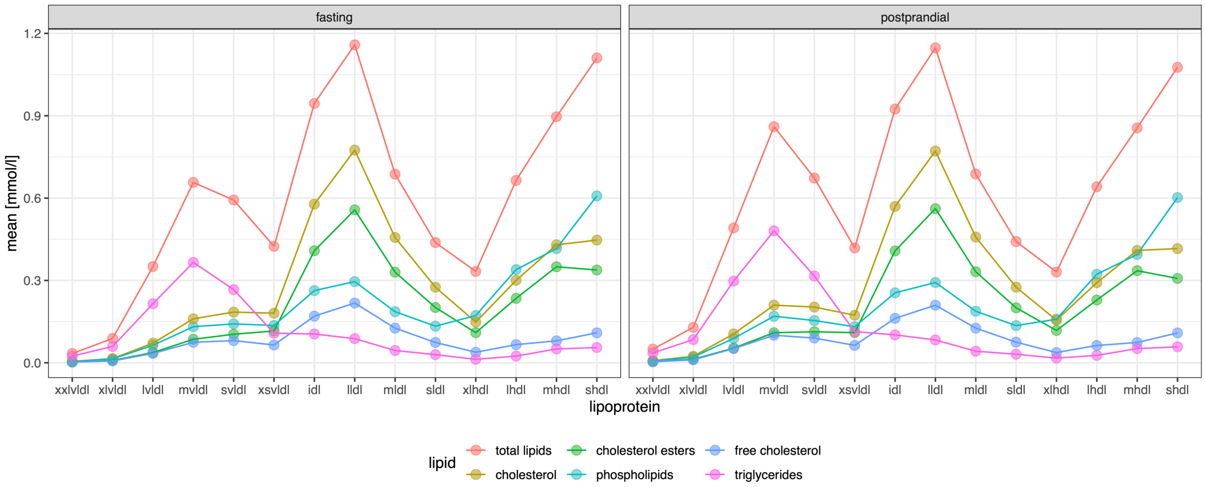

**S4 Figure: Mean lipid estimates across lipoproteins.** For each lipoprotein (x-axis), organized by density or size (most dense to least dense – left to right) the mean (units = mmol/l) of each lipid fraction was estimated and plotted (y-axis). Each lipid fraction is plotted in a different color as indicated in the legend.

### S5 Figure: Postprandial-Fasting delta values

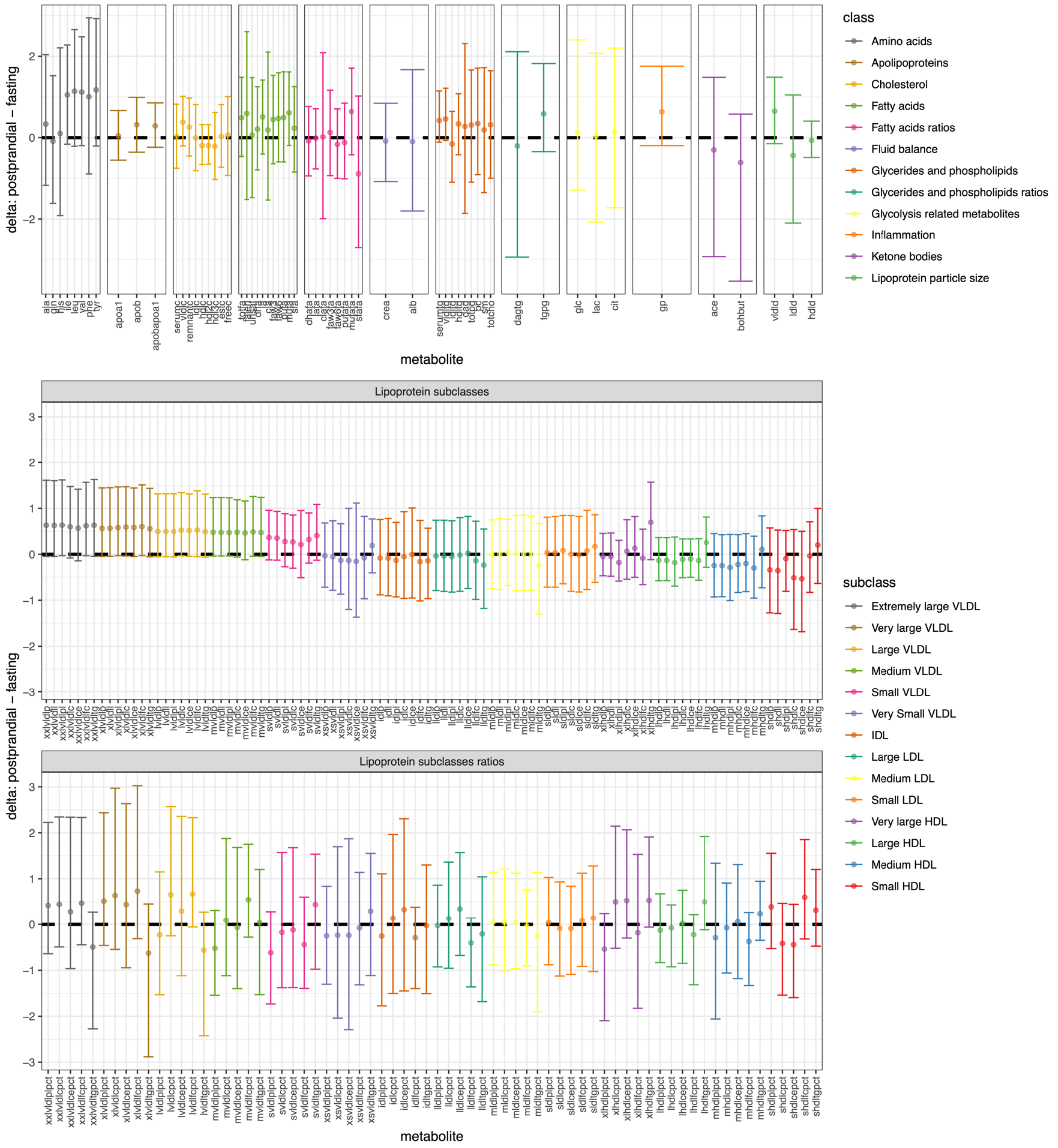

**S5 Figure: Postprandial-Fasting delta value.** A simple delta value (postprandial - fasting) was estimated for each sample by metabolite, after centring (mean=0) and scaling (SD=1) the fasting and postprandial data together. Plotted here, for each metabolite (x-axis) is the mean delta (point) and the 95% confidence intervals (whiskers) of the delta distribution (y-axis). The y-axis are on a fixed scale across all three plot rows. The top row illustrates values for metabolites partitioned and coloured by their respective class. The middle and bottom row illustrate the lipoproteins and lipoprotein ratios, respectively, coloured by their subclass or lipoproteins – organized from least dense to most dense, left to right.

### S6 Figure: Observational effect estimates by dietary state and class

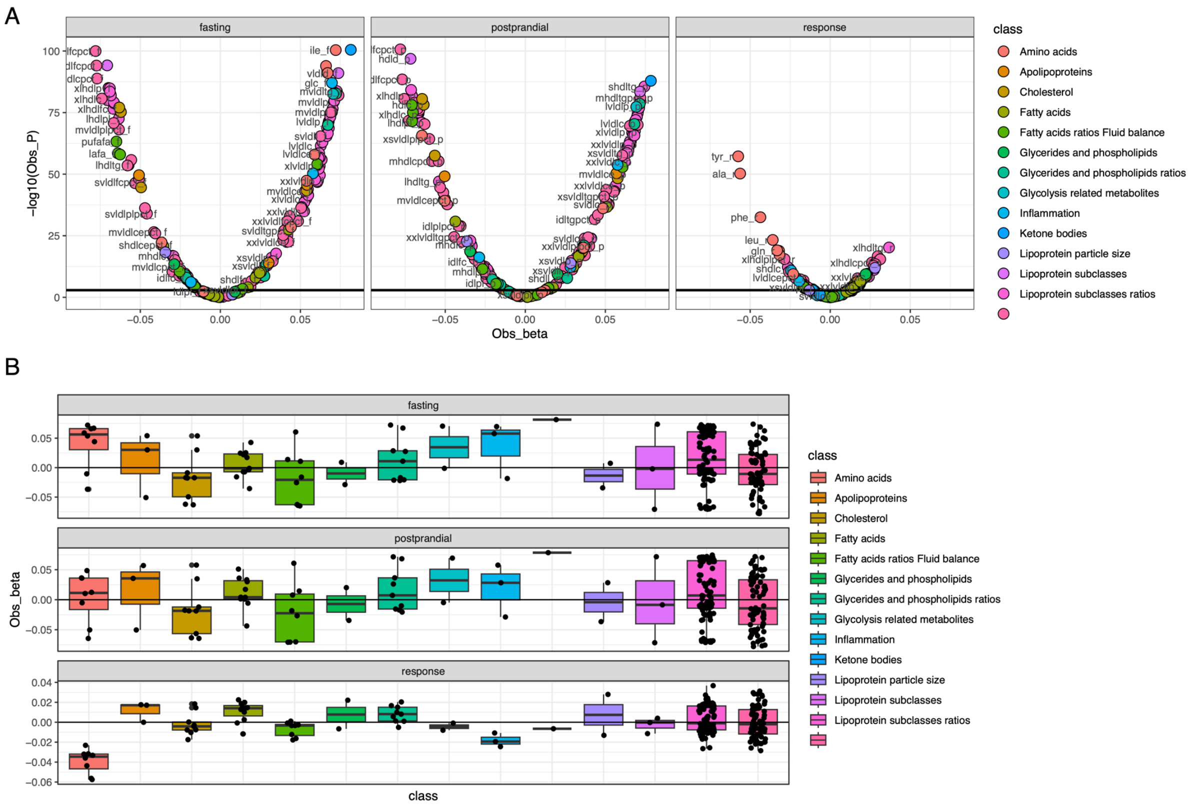

**S6 Figure: Observational effect estimates by dietary state and class. (A)** BMI – metabolite effect estimates and their relationship with the linear model P-value (-log10(P)) are illustrated in the volcano plots, one for each dietary state (fasting, postprandial, and response). Along the x-axis are the point or effect estimate and along the y-axis are the -log10(P-values). Metabolites are classified and colored by Nightingale Health class assignments. The alpha threshold for defining a metabolite as associated with BMI is indicated by the horizontal black line (P<0.05/40). **(B)** BMI – metabolite observational effect estimates are classified by dietary state (fasting, postprandial and response) and Nightingale Health class assignments (x-axis and color key) to illustrate the distribution and mean effect for each class of metabolite by dietary state. Each box defines the mean (box horizontal line), the 25$^{th}$ and 75$^{th}$ percentile of the data distribution (limits of the box), and the 1.5 * inter-quartile range of the box (whiskers) as defined by the ggplot function geom_boxplot().

#### S7 Figure: Variance explained in BMI by associated covariables

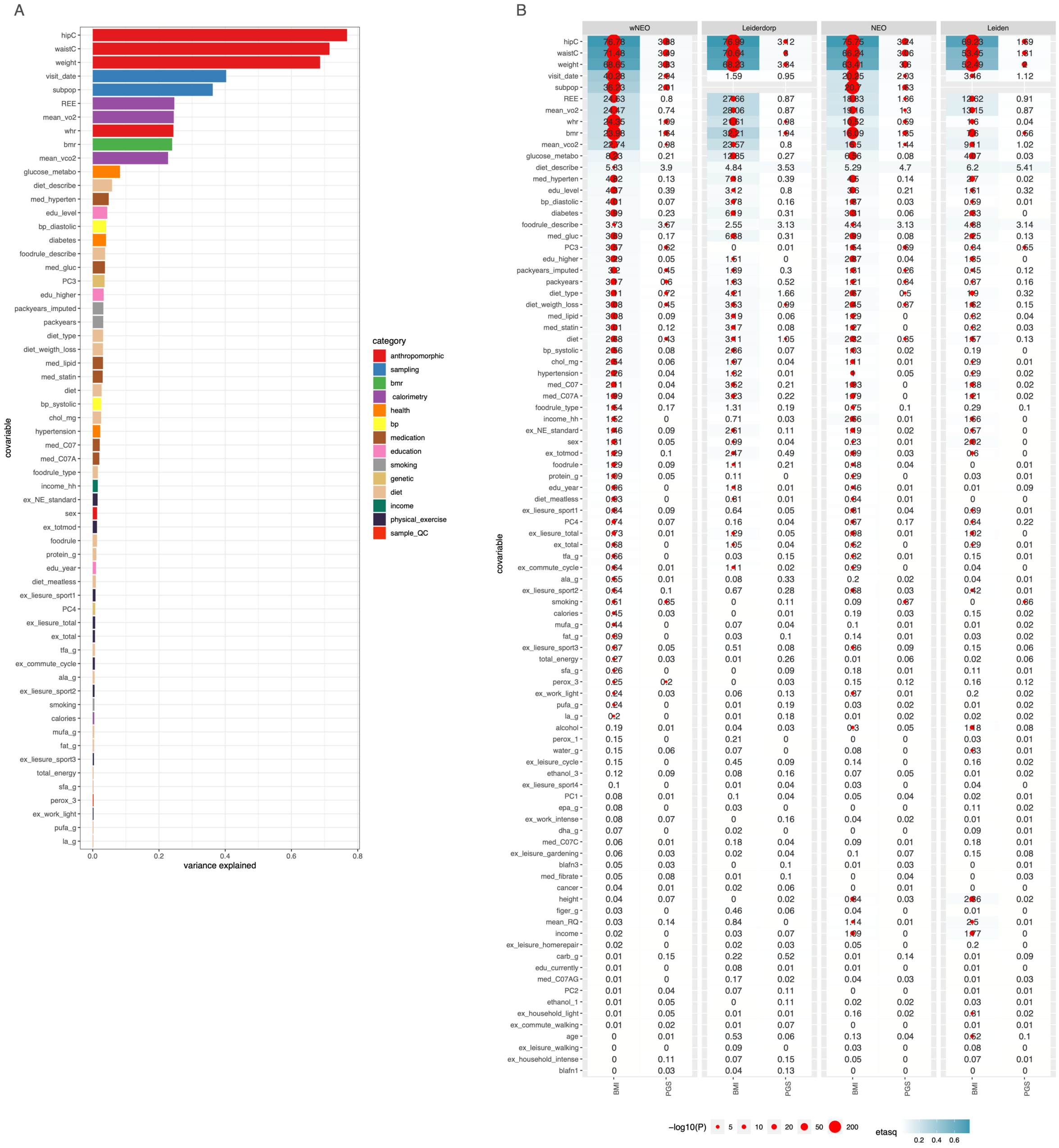

**S7 Figure: Variance explained in BMI by associated study covariables.** The x-axis provides an (weighted; wNEO) estimate of the variance explained (eta-squared) in BMI by each study covariable as derived by extracting the sums of squares from an analysis of variance. The y-axis provides the name of all covariables that associate with BMI. Covariable names can be mapped to descriptions in Supplementary Table 2. Study covariables were placed into categories (color key) and used to color each bar, which are also available in Supplementary Table 2. Plot values can be found in Supplementary Table 4. **(B)** For each (sub-)population (plot columns) analysis (1) wNEO, (2) Leiderdorp, (3) NEO, and (4) Leiden univariable linear models and analysis of variances (ANOVA) were run with either BMI or BMI-PGS (x-axis) set as the dependent and each covariable (y-axis) was defined as the independent. From each ANOVA the sums of squares were extracted, and an estimate of the variance explained in the dependent was derived in the form of an eta-squared ($\eta^2$) statistic (blue shading and cell text ($\eta^2$*100)). For each analysis where an association was observed ($P<0.05/40$) a red dot (scaled by -log10(ANOVA F-test P)) can be found in the relevant cell. The y-axis is ordered by the wNEO BMI $\eta^2$.

### S8 Figure: Variance explained in PGS by associated covariable

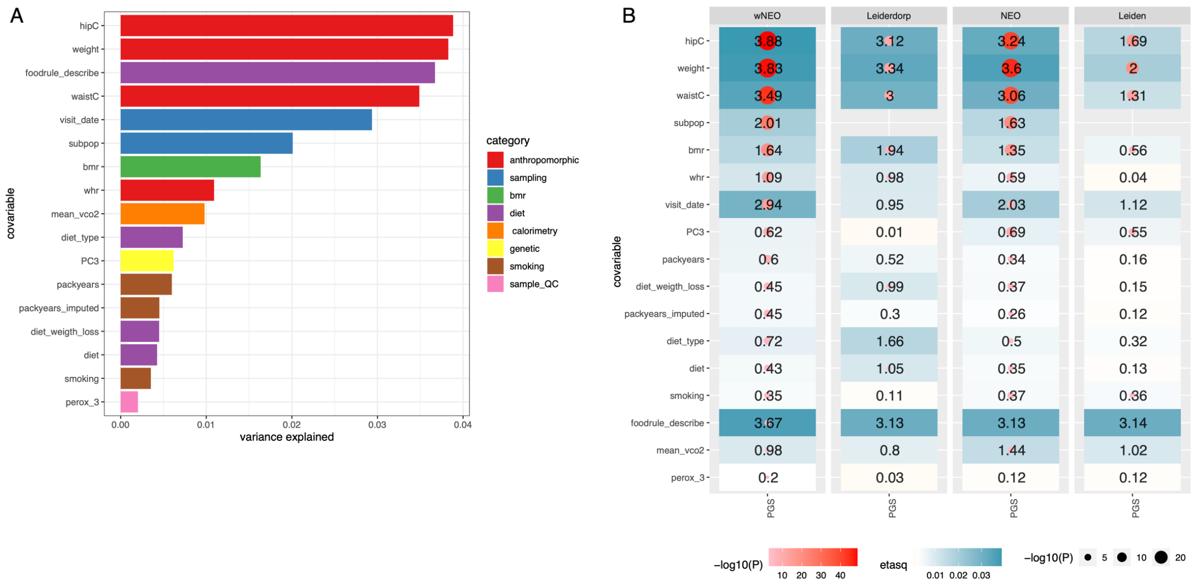

**S8 Figure: Variance explained in BMI-PGS by associated study covariables. (A)** The x-axis provides an (weighted; wNEO) estimate of the variance explained (eta-squared) in BMI-PGS by each study covariable as derived by extracting the sums of squares from an analysis of variance. The y-axis provides the name of all covariables that associate with BMI. Covariable names can be mapped to descriptions in Supplementary Table 2. Study covariables were placed into categories (color key) and used to color each bar, which are also available in Supplementary Table 2. Plot values can be found in Supplementary Table 4. **(B)** For each (sub-)population (plot columns) analysis (1) wNEO, (2) Leiderdorp, (3) NEO, and (4) Leiden univariable linear models and analysis of variances (ANOVA) were run with BMI-PGS (x-axis) set as the dependent and each covariable (y-axis) associated with BMI-PGS in the wNEO analysis was defined as the independent. From each ANOVA the sums of squares were extracted, and an estimate of the variance explained in the dependent was derived in the form of an eta-squared ($\eta^2$) statistic (blue shading and cell text ($\eta^2*100$)). For each analysis where an association was observed ($P<0.05/40$) a red dot (scaled by -log10(ANOVA F-test P)) can be found in the relevant cell. The y-axis is ordered by the wNEO p-value.

### S9 Figure: Metabolite associations with “BMI-PGS associated” covariables

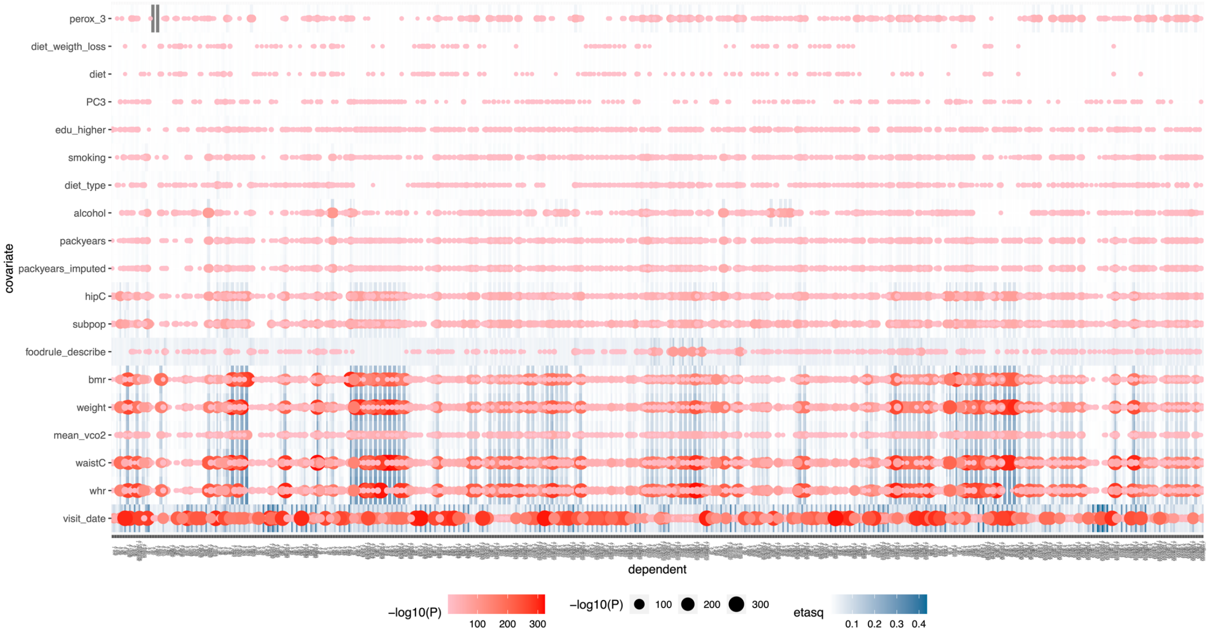

**S9 Figure: Metabolite associations with “BMI-PGS associated” covariables.** For each metabolite (x-axis) a univariable linear models and analysis of variances (ANOVA) were run against each covariable (plus alcohol and higher education) previously observed to be associated with BMI-PGS in univariable analyses (y-axis). Each metabolite trait was set as the dependent and each covariable was set as the independent variable. From each ANOVA the sums of squares were extracted, and an estimate of the variance explained in the dependent was derived in the form of an eta-squared ($\eta^2$) statistic (blue shading). For each analysis where an association was observed ($P<0.05/40$) a red dot (scaled by -log10(ANOVA F-test P)) can be found in the relevant cell. The y-axis is ordered by the average $\eta^2$ across all metabolites.

#### S10 Figure: MR effect estimates by dietary state and class

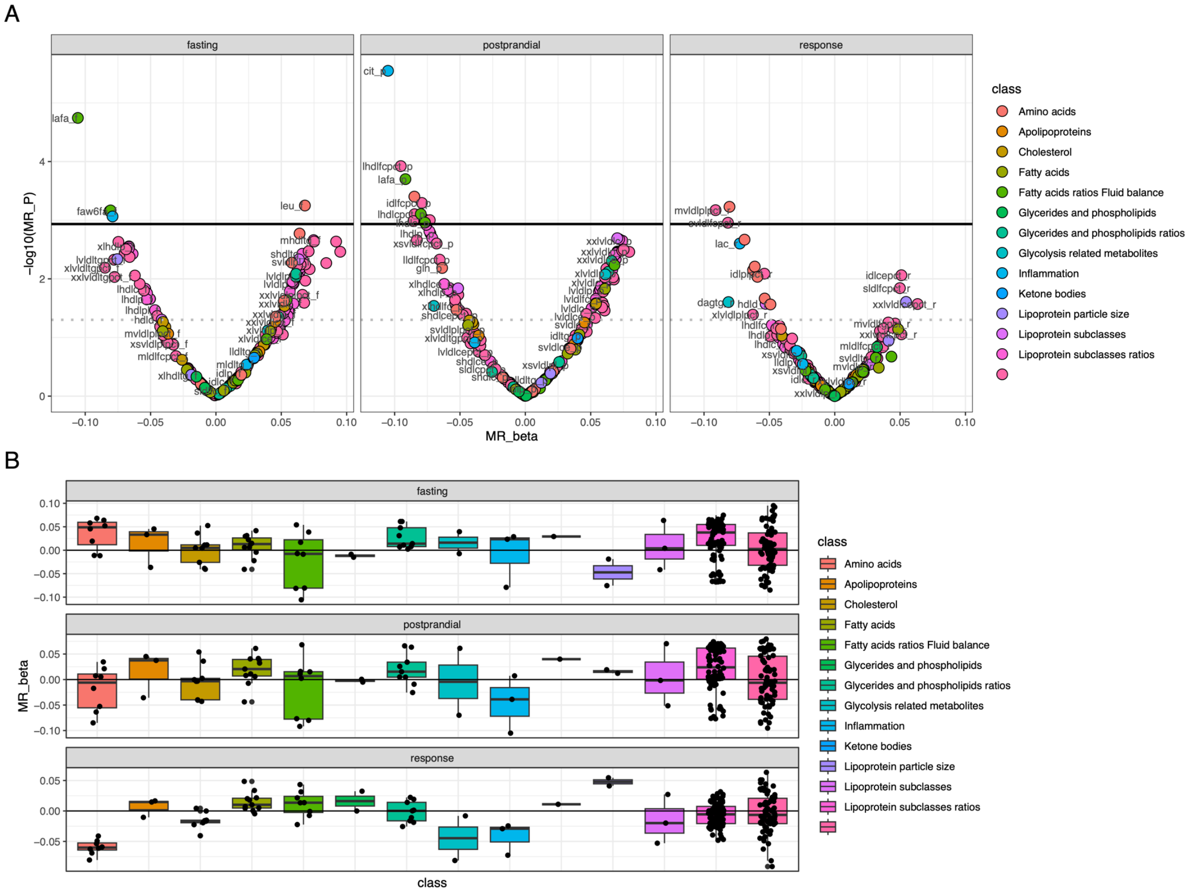

**S10 Figure: MR effect estimates by dietary state and class. (A)** BMI – metabolite MR effect estimates and their relationship with the linear model P-value (-log10(P)) are illustrated in the volcano plots, one for each dietary state (response, postprandial and fasting). Along the x-axis are the point or effect estimate and along the y-axis are the -log10(P-values). Metabolites are classified and colored by Nightingale Health class assignments. The alpha threshold for defining a metabolite as nominally associated (P<0.05) is the dotted grey line, and associated with BMI is indicated by the horizontal black line (P<0.05/43). **(B)** BMI – metabolite MR effect estimates are classified by dietary state (response, postprandial, and fasting) and Nightingale Health class assignments (x-axis and color key) to illustrate the distribution and mean effect for each class of metabolite by dietary state. Each box defines the mean (box horizontal line), the $25^{th}$ and $75^{th}$ percentile of the data distribution (limits of the box), and the 1.5 * inter-quartile range of the box (whiskers) as defined by the ggplot function geom_boxplot().

#### S11 Figure: Scatter plot of Obs. & MR effect estimates by sample population

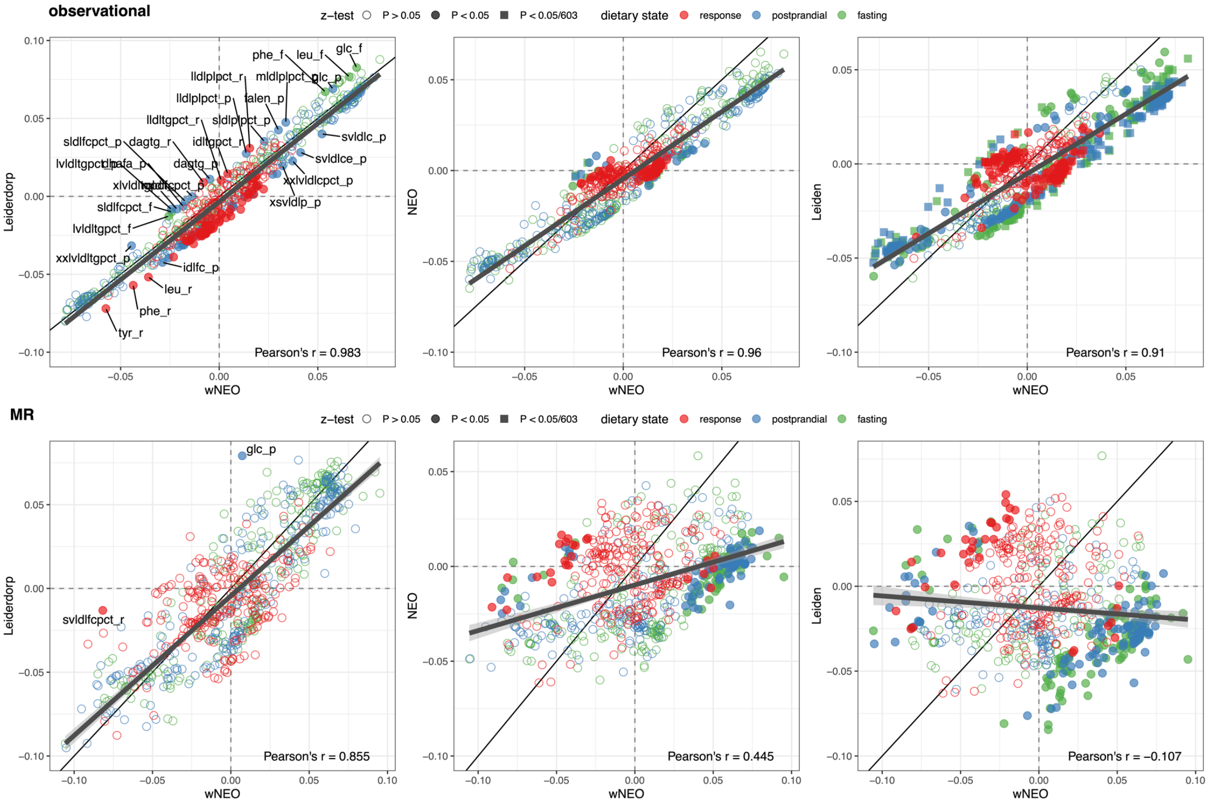

**S11 Figure: Scatter plot of Obs. & MR effect estimates by(sub)-population.** Observational (top) and MR (bottom) effect estimates are compared between the weighted NEO (wNEO; x-axis) analyses and each other sample (sub)-population (y-axis) – left: Leiderdorp, middle: NEO, right: Leiden. Plotting dots in red, blue, and green denote effect estimates for metabolites in the response, postprandial, and fasting dietary states, respectively. Solid circles represent effect estimates that differ, as test by a z-test, between the two (sub-)sample populations at a p-value of 0.05. Solid squares are those that differ at a Bonferroni adjusted p-value of 0.05/603. The thin black line represents an equivalency line (intercept = 0, slope = 1) and the thick black line is the best fit line through the data. An estimate of Pearson's r correlation coefficient is in the lower right corner of each plot.

### S12 Figure: NEO and Wurtz et al MR estimate scatter plot

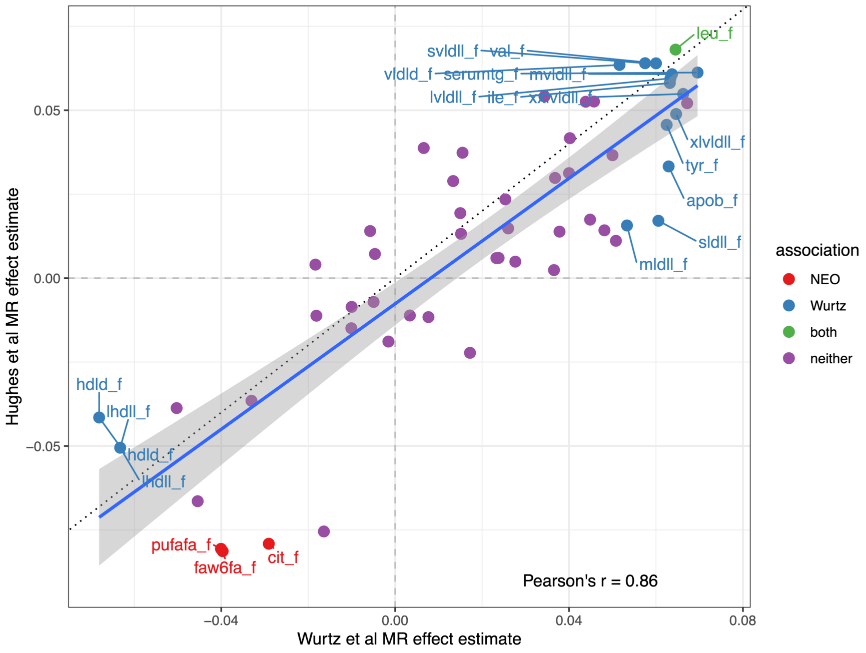

**S12 Figure: NEO and Wurtz et al MR estimate scatter plot.** Each of the 57 fasting, matched, metabolites are represented by a point. The x-axis provides the MR effect estimate as observed by Wurtz et al, and the y-axis is the MR effect estimate observed in this study (wNEO framework). The dotted black line is an equivalency line. The blue line is the best fit regression line. Estimates that were defined as associated in both NEO and Wurtz et al are colored green, those specific to NEO are colored red, those specific to Wurtz et al are colored blue, and those with no MR association in either study are purple. The associated metabolites are labelled with their metabolite ID in the plot. See Supplementary Table 1 for a further annotation.

### S13 Figure: Postprandial observational lipoprotein profile

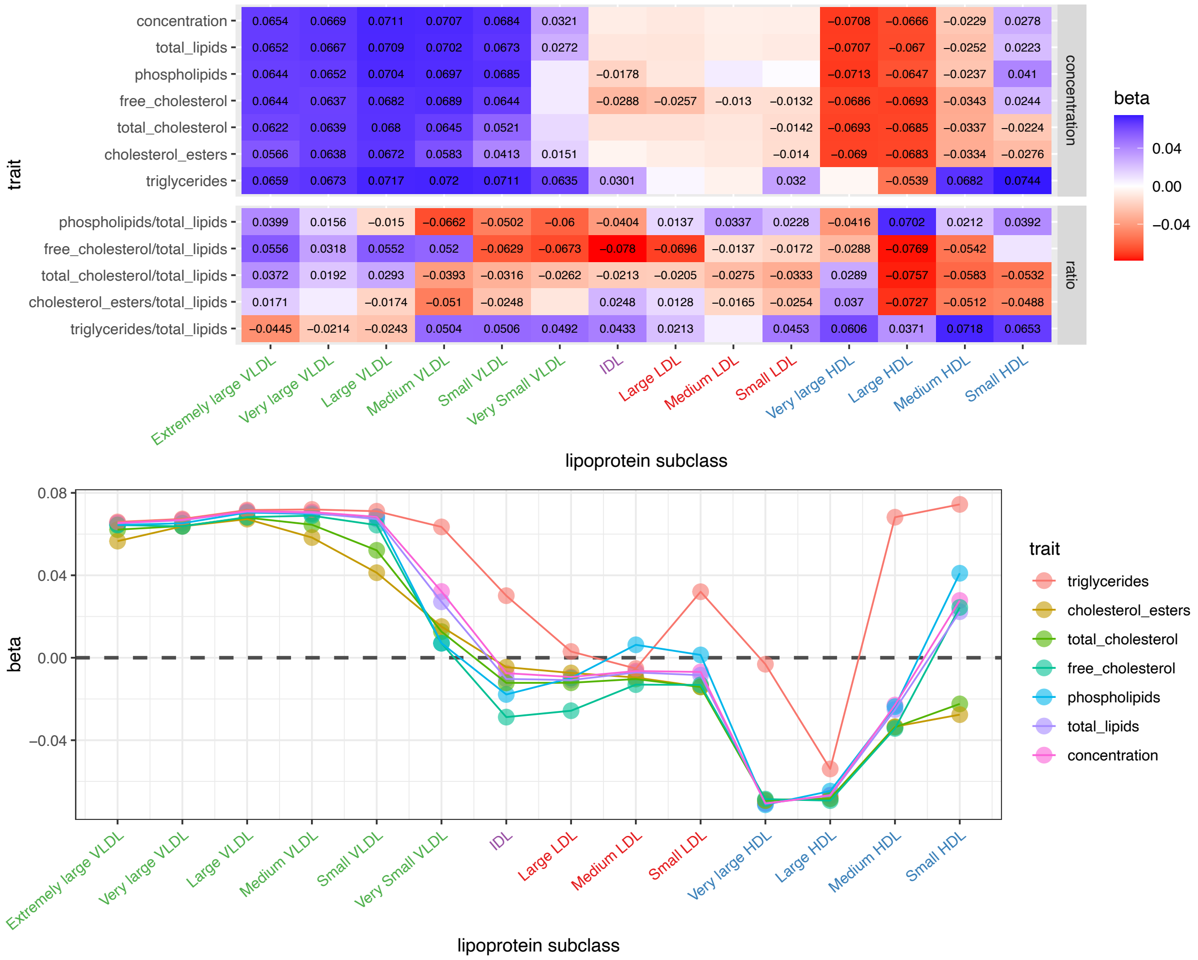

**S13 Figure: Postprandial observational profile.** *Upper*: A tile plot of observational effect estimates for lipoproteins and lipoprotein ratios in the postprandial state. Tiles with an effect estimate provided in text are those with a p-value smaller than 0.05. The lipoproteins are labeled and color corradiated the x-axis, and the component or ratio being measured is along the y-axis. *Lower*: A dot plot or profile of postprandial observational effect estimates for lipoproteins (x-axis) ordered by lipoprotein size or density is provided to illustrate the correlation between effect estimates (y-axis) within a lipoprotein and the structure of estimates between lipoproteins by size. The component or measurement of each lipoprotein are defined by the color as described in the key.

#### S14 Figure A: Fasting MR lipoprotein profile

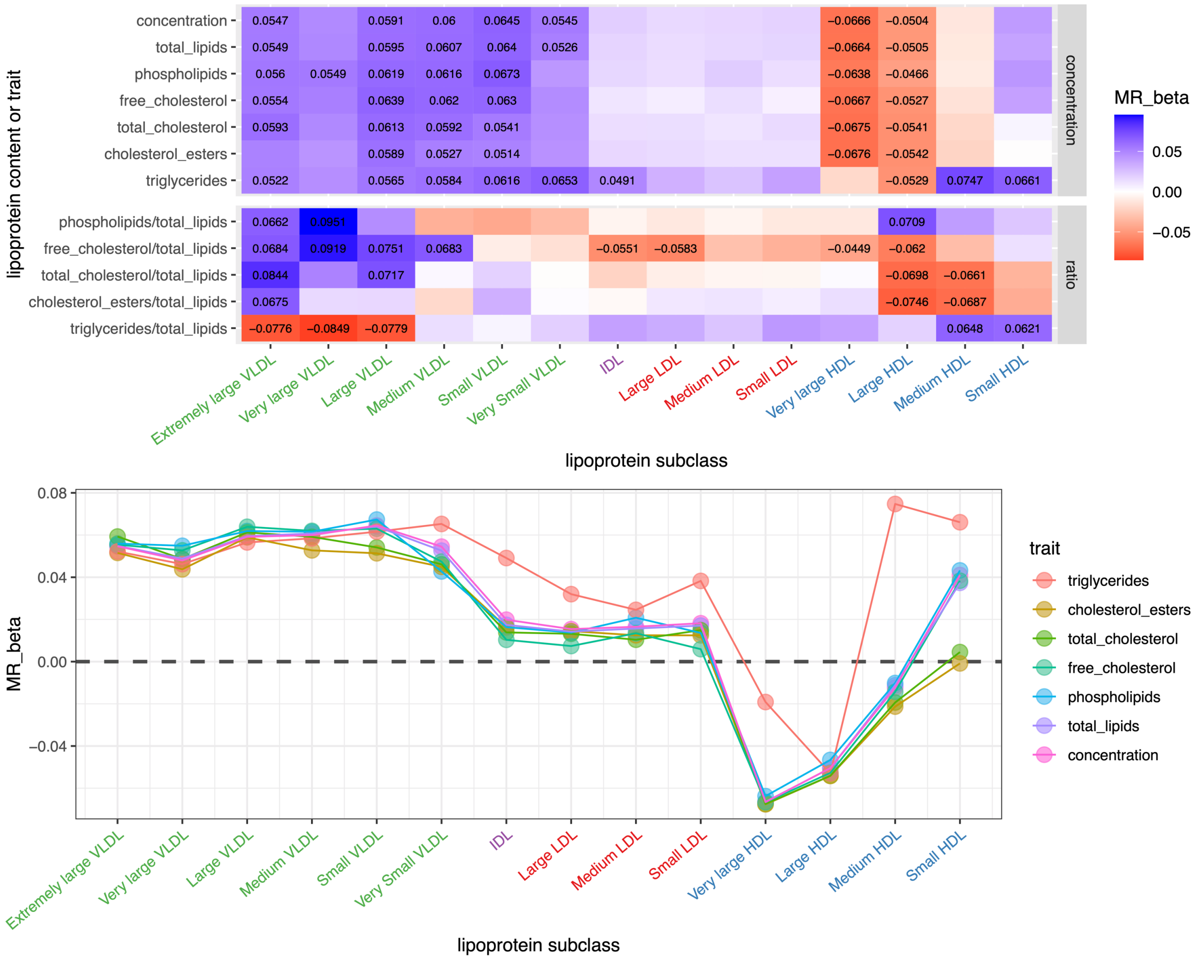

**S14 Figure A: Fasting MR profile.** *Upper*: A tile plot of MR effect estimates for lipoproteins and lipoprotein ratios in the fasting state. Tiles with an effect estimate provided in text are those with a p-value smaller than 0.05. The lipoproteins are labeled and color corradiated the x-axis, and the component or ratio being measured is along the y-axis. *Lower*: A dot plot or profile of fasting MR effect estimates for lipoproteins (x-axis) ordered by lipoprotein size or density is provided to illustrate the correlation between effect estimates (y-axis) within a lipoprotein and the structure of estimates between lipoproteins by size. The component or measurement of each lipoprotein are defined by the color as described in the key.

### S14 Figure B: Postprandial MR lipoprotein profile

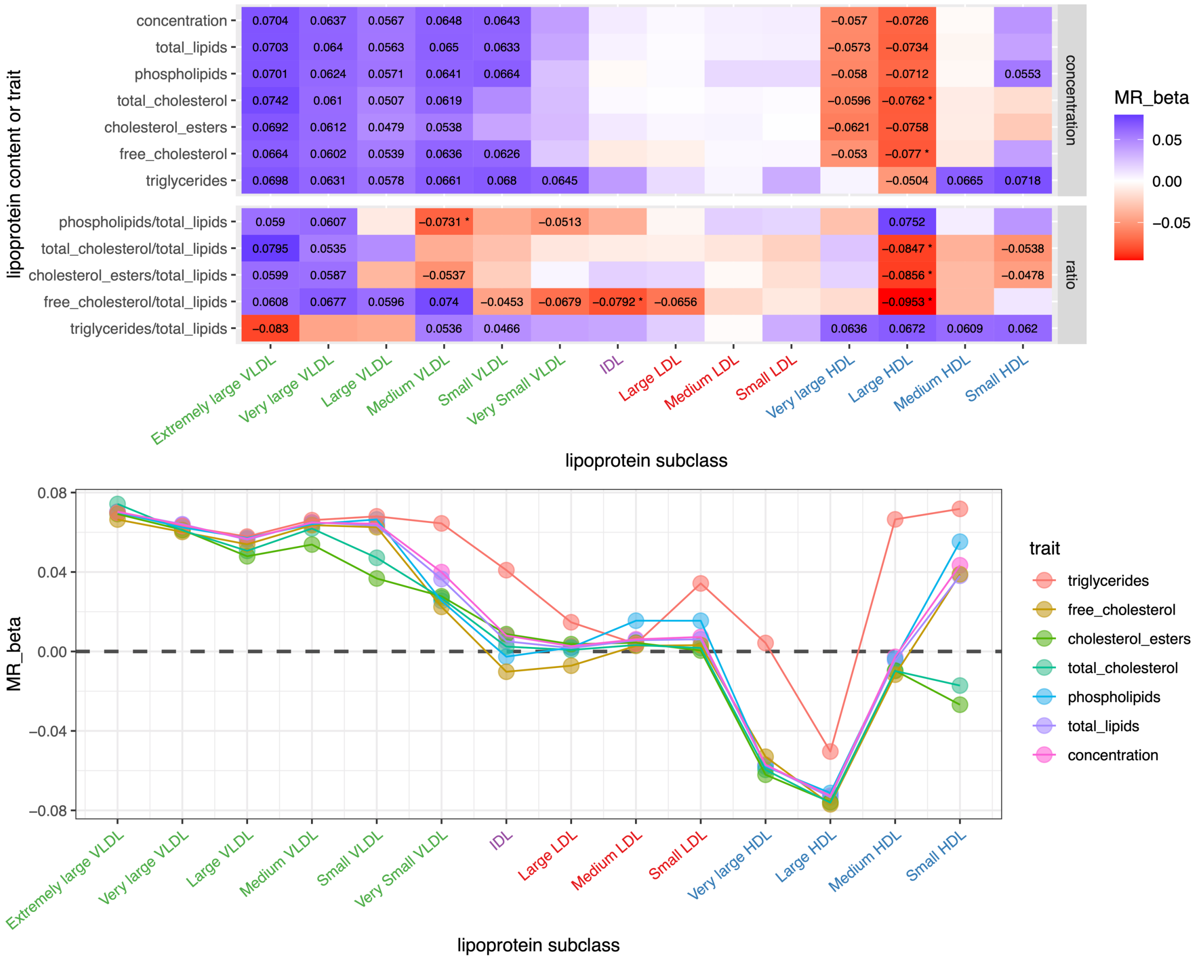

**S14 Figure B: Postprandial MR profile.** *Upper*: A tile plot of MR effect estimates for lipoproteins and lipoprotein ratios in the postprandial state. Tiles with an effect estimate provided in text are those with a p-value smaller than 0.05. The lipoproteins are labeled and color corradiated the x-axis, and the component or ratio being measured is along the y-axis. *Lower*: A dot plot or profile of fasting MR effect estimates for lipoproteins (x-axis) ordered by lipoprotein size or density is provided to illustrate the correlation between effect estimates (y-axis) within a lipoprotein and the structure of estimates between lipoproteins by size. The component or measurement of each lipoprotein are defined by the color as described in the key.
